# Normative EEG Effective Connectivity as a neural marker of Autism Spectrum Disorder

**DOI:** 10.64898/2026.07.27.26359031

**Authors:** Lorenzo Gaetano Amato, Federico Fattorini, Alberto Arturo Vergani, Marco Pagani, Nicolò Meneghetti, Alberto Mazzoni, Michelangelo Fabbrizzi

**Author notes:** These authors gave equal first author contributions (Lorenzo Gaetano Amato, Federico Fattorini). These authors gave equal senior author contributions (Alberto Mazzoni, Michelangelo Fabbrizzi).

## Abstract

Autism spectrum disorder (ASD) is one of the most common neurodevelopmental disorders. The absence of a clear aetiology strengthens the need to identify quantitative biomarkers capable of diagnosing the condition and assessing the severity of symptoms. We implemented a normative modelling framework based on EEG effective connectivity (EC), leveraging recordings collected in three centers, in typical development (TD) and ASD participants with age spanning from 5 to 19 years. Deviations from the normative evolution were tested as ASD biomarkers, compared with standard EC metrics. Normative EC metrics supported accurate classification of ASD vs TD participants (0.82 AUC), reaching 0.95 AUC when combined with behavioral subscales, significantly outperforming a classification based on behavioral data alone. In ASD participants, selected metrics in the Temporo-Parietal-Frontal network predicted social responsiveness scale (SRS) values with high significance (SRS Total *R*^2^=0.42, p=0.0005). These results suggest that normative EC values constitute robust and generalizable ASD biomarkers.

## Introduction

Autism spectrum disorder^1,2^ (ASD) has lately emerged as one of the most common neurodevelopmental disorders. Current reports show that 1% of the world population lives with ASD^3,4^. Incidence is higher in developed countries, reaching 3.2% in children aged between 4 and 8 years^5^.A key challenge in current ASD research is the identification of robust and translational markers of the condition, able to support a precise diagnosis of the condition in different socioeconomic backgrounds^6,7^ without requiring expensive machinery.

Emerging biomarkers derived from quantitative medical exams^8^ such as genetic^9,10^, neuroimaging^11–13^, metabolic^14^ and neurophysiological^15^ have shown promise for ASD diagnosis. However, multi- center validation and prospective studies remain lacking^8^, casting doubt on the translational capabilities of these methodologies. The few studies that reported multi-center generalization reported marked drop in diagnostic accuracy when generalizing results^11^.

Currently, ASD diagnosis is primarily based on behavioural and cognitive scales^16^ such as the Autism Diagnostic Observation Schedule-Second Edition and the Autism Diagnostic Interview. These quantitative metrics present high but not decisive classification accuracies, with sensitivity of 91% and 80% and specificity of 76% and 72%, respectively^17^. Other methods such as the Social Responsiveness Scale^18^ (SRS), present good generalizability in homogenous cultural contexts^19^ but are prone to high variability in terms of age, cognitive level and expressive language^20^.

Among candidate markers for ASD diagnosis, EEG has emerged as a scalable, equitable and cost-effective tool^21^. Quantitative EEG studies have investigated several possible neural markers related to ASD pathophysiology, including spectral^22^, complexity^23^, and both functional^24^ and effective connectivity^25,26^ metrics. EEG studies previously attempted ASD classification and cross-modal regression of behavioural manifestations of the condition^15^. However, there is no current EEG or EC metric capable of simultaneously supporting a robust ASD identification and tracking the severity of symptoms.

A novel introduction in ASD research has been that of normative modelling^27–31^, allowing researchers to investigate age-dependent deviations of ASD subjects in terms of behavioural or quantitative clinical quantities. The relevance of considering divergence from normative developmental trajectories as a marker of ASD was emphasized by Lombardo et al.^32^, who highlighted its contribution to distinguishing ASD subtypes. At the same time, Lin et al.^33^ questioned the existence of discrete subtypes, while still underscoring variability from typical development as a core dimension of ASD. Recent advancements in ASD normative modeling were described by Mandelli et al.^29^, combining ASD subtyping with normative modeling of behavioral scales to enhance prognostic prediction. In ASD, normative modeling has been applied to sMRI recordings by Zabihi et al.^30^, and Betlehem et al.^31^ focusing on cortical thickness evolution along early lifespan, and in terms of fMRI networks by Jiang et al.^34^. However, normative modeling of EEG metrics for ASD research has never been reported to the best of our knowledge.

Here, we present a normative modelling pipeline (Figure 1), investigating the normative evolution of EEG across early age lifespan (5-19 years). All ASD and TD subjects were part of the publicly available Healthy Brain Network (HBN) dataset^35^. Normative evolution was obtained from Typically Developing (TD) subjects, acquired across two centers. Normative EC values from ASD participants (acquired in three centers) were then analyzed. Normative EC metrics supported an accurate classification of TD vs ASD participants across centers. In combination with SRS metrics, they significantly improved ASD identification based on behavioral metrics. Furthermore, they were used to predict total SRS values, identifying selective EC metrics related to the Temporo-Parietal-Frontal network which robustly predicted behavioural SRS subscales. Normative EC metrics emerged as robust and cost-effective metrics, capable of simultaneously discriminating ASD vs TD and relating to the severity of behavioural manifestations of the condition.

**Figure 1.**
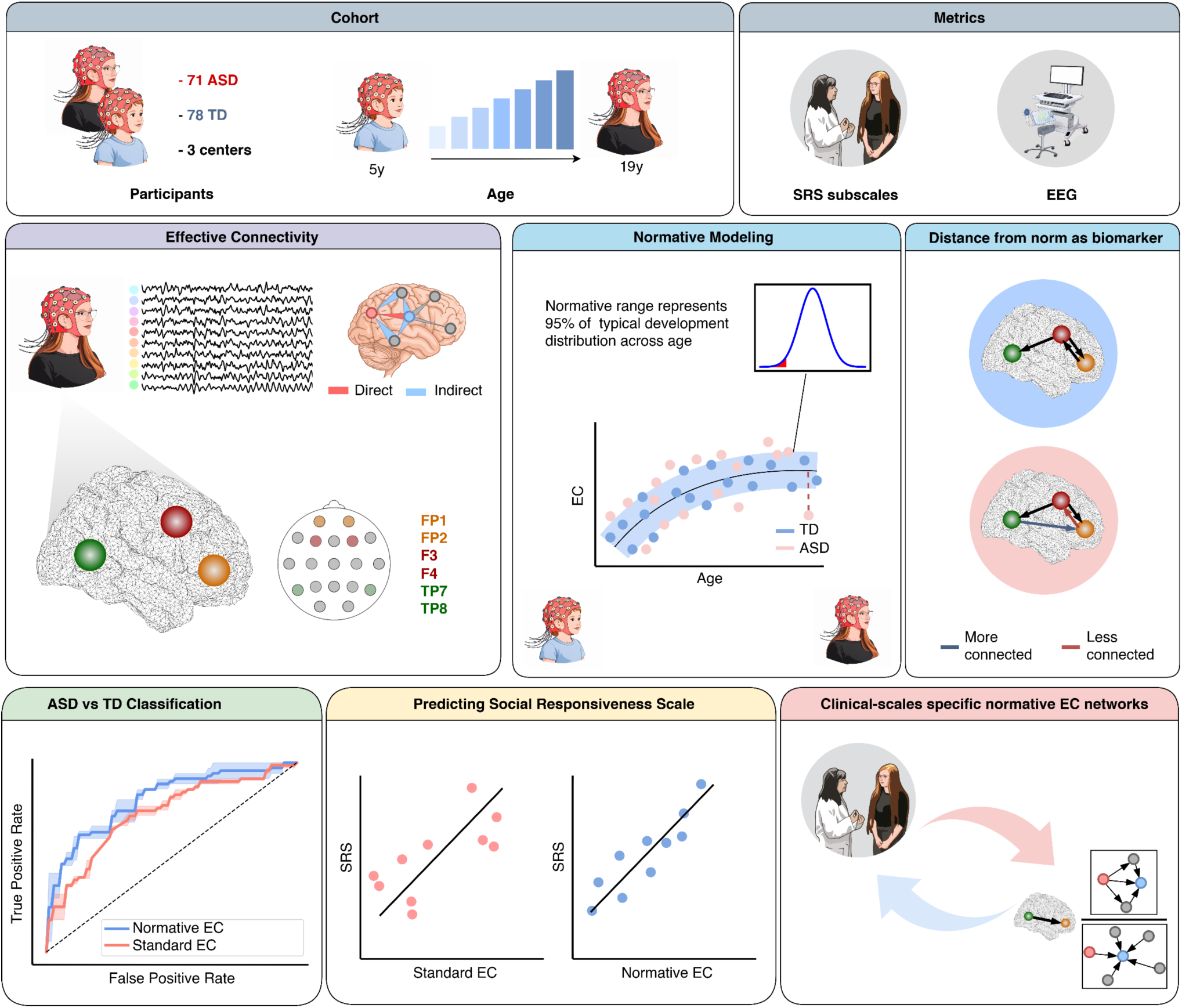
Normative Effective Connectivity pipeline: 78 TD participants and 71 ASD from three different centers from database HBN were considered in this study. The age of participants spanned 5-19 years. SRS values and resting-state EEG recordings were analyzed for all participants. Effective connectivity values between Frontal and Temporo-Parietal channels were computed from EEG recordings, extracting different metrics, each measuring different aspects of neural connectivity. Normative modeling was then derived from TD metrics, and variability from the norm was evaluated as an ASD biomarker. Evaluation of EC metrics comprised: (i) ASD vs TD classification, (ii) prediction of SRS values and (iii) identification of normative EC metrics and networks selectively associated with SRS values.

## Results

EEG recordings of 78 TD and 71 ASD participants from the HBN dataset (see Methods) were analyzed. Data were collected from three centers (Supplementary Table 1). 9 TD and 13 ASD were removed after preprocessing due to EEG data cleaning and following noisy channel removal (see Methods), while SRS values were not available for 1 TD and 1 ASD subject, leading the final number to 68 TD and 57 ASD participants. No statistical differences were found between groups in terms of age and sex composition (Supplementary Table 2). SRS subscales were evaluated for both groups, with TD subjects showing significantly lower values (U=434, p<0.00001 for SRS Total, Supplementary Table 2). Different clinical centers presented no significant interaction with demographic and SRS differences (Supplementary Table 2).

### Normative Effective Connectivity discriminates between Autism Spectrum Disorder and Typically Developing

EEG recordings were then analyzed to compute effective connectivity metrics. Frontal channels (Fp1, Fp2, F3 and F4) and Temporo-Parietal channels (Tp7 and Tp8) were included in the EC computation. Sixteen different EC connectivity metrics, sensitive to different connectivity motifs, were measured. These metrics included Coherence, Spectral Density, Granger Geweke Causality, Partial Directed Coherence (PDC) and Direct Transfer Function (DTF) in different formulations ( see Delorme et al.^36^ for details on the different metrics). All employed metrics are summarized in Table 1.

**Table 1.** Description of Effective Connectivity metrics employed in the study. For reference we refer to the SIFT Toolbox documentation page (https://github.com/sccn/SIFT/blob/master/documentation/).

|  | Family | Description |
| --- | --- | --- |
| **Spectral Density Matrix (S)** | Spectral Entropy | Cross-spectral coupling |
| **Coherence (Coh)** | Coherence | Frequency-domain equivalent of cross-correlation |
| **Imaginary part of Coh (iCoh)** |  | By considering only the Imaginary part of coherence the measure is invariant to zero-lag connections |
| **Partial Coh (pCoh)** |  | Conditional coherence of a channel with respect to other measured channels |
| **Multiple Coh (mCoh)** |  | Univariate quantity evaluating total coherence of a channel with all other measured channels |
| **Granger-Geweke Causality (GGC)** | Granger Causality | Granger causality in the Geweke formulation |
| **Partial Directed Coherence (PDC)** | PDC | Conditional GC between two electrodes normalized by the total amount of causal connections of the input channel |
| **Normalized PDC (nPDC)** |  | Conditional GC between two electrodes normalized with a noise-robust denominator |
| **Generalized PDC (GPDC)** |  | Conditional GC between two electrodes with a different formula using residuals to normalize |
| **PDC Factor (PDCF)** |  | Conditional GC between two electrodes normalized by the total amount of causal connections of the target channel |
| **Renormalized PDC (RPDC)** |  | Conditional GC between two electrodes normalized by the total power of the target channel |
| **Direct Transfer Function (DTF)** | DTF | Total information flow between two channels. |
| **Normalized DTF (nDTF)** |  | Total information flow between two channels normalized by the total amount of connections of the target channel |
| **Full-Frequency DTF (ffDTF)** |  | Total information flow between two channels normalized by the total amount of connections of the target channel in a frequency-independent manner |
| **Directed DTF (dDTF)** |  | Product between ffDTF and pCoh |
| **Thresholded dDTF (dDTF08)** |  | Product between ffDTF and pCoh with a significance threshold. |

Band-average EC metrics were computed in standard EEG bands (delta 0.5-4Hz, theta 4-8Hz, alpha 8-13Hz, beta 13-30Hz, gamma 30-45 Hz, see Methods) for all participants (Figure 2a). Subsequently, Normative Modeling evolution was computed for all metrics from TD subjects, obtaining normative curves for age-related evolution (see Methods). For all subjects, normative EC metrics were then computed as the individual deviation from the norm (Figure 2b). For both standard and normative EC metrics, correlation matrices were computed to assess similarity in different metrics employed (Supplementary Figure 1). Both standard and normative EC metrics presented low correlation across metrics: r=0.095±0.093 and r=0.082±0.070, respectively. Correlation matrix of standard EC metrics presented no significant differences with respect to the correlation matrix of normative EC metrics (Mann-Whitney U-test=23424, p=0.17).

**Figure 2:**
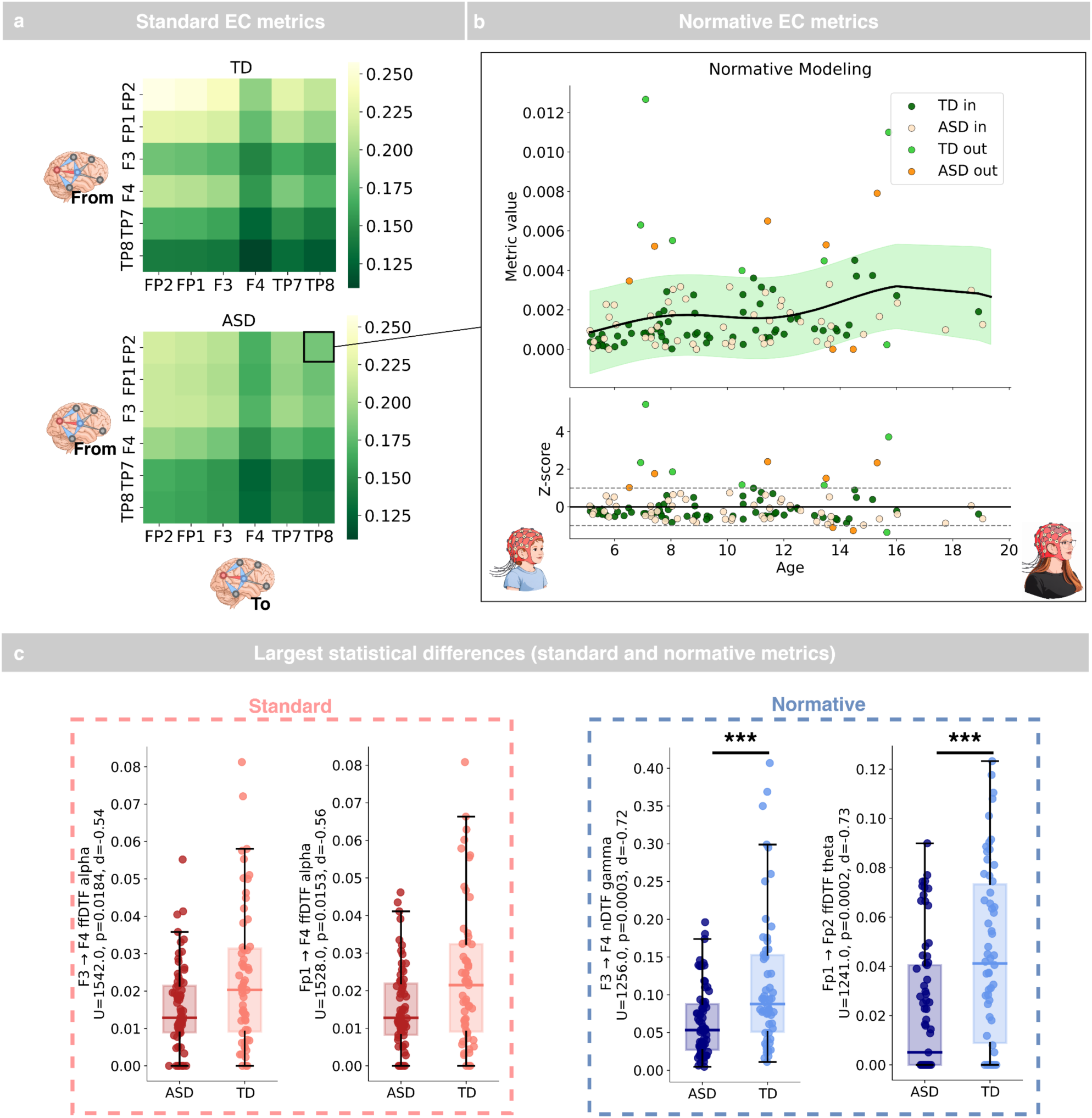
Statistical differences between normative and standard EC metrics. **(a):** Standard EC metrics were computed for each TD and ASD participants, the panel shows an example of metric/subject (dDTF08) where each cell represents the band average value of a directional connection from an electrode to another. **(b):** Normative curves of EC were obtained from TD data. Deviation from the norm was then computed for each participant, and checked as an ASD biomarker. As an example, it is reported the dDTF08 TP8→Fp2 in the Beta band. The shaded area represents 95% confidence intervals of the normative model. **(c):** Comparison of the standard EC metrics and normative EC metrics, with the best two standard EC metrics (ffDTF from F3 to F4 in alpha band and ffDTF from Fp1 to F4 in alpha band,left) and the best two normative EC metrics (nDTF from F3 to F4 in gamma band and ffDTF from Fp1 to Fp2 in theta band, right) highlighted. Each point of the box-plot represents a participant. Horizontal lines represent median values, while whiskers are set at 1.5 times the interquartile range. Significance notation: *** stands for p<0.0005, p-values are reported after false discovery rate correction.

Among standard EC, only 10 metrics presented a large effect size (Cohen’s d>0.5) in discriminating between ASD and TD subjects, while 52 metrics presented a large effect size for normative EC metrics. The top 2 metrics (defined as those that presented the highest effect sizes in terms of the ASD vs TD differences, Figure 2c) for Standard EC metrics were the full frequency DTF (ffDTF) Fp1-F4 in alpha band (U=1528.0, p=0.015, d=0.56) and ffDTF F3→F4 in alpha band (U=1542, p=0.018, d=0.54). The top 2 normative EC metrics were the ffDTF Fp1→Fp2 in the theta band (U=1241.0, p=0.0002, d=0.73) and the normalized DTF (nDTF) F3→F4 in gamma band (U=1256.0, p=0.0003, d=0.72).

### Normative Effective Connectivity supports robust ASD vs TD classification

Normative EC metrics were evaluated for ASD versus TD classification using a nested cross-validation framework (Figure 3a). Classification was performed using a logistic regression classifier with L2 regularization, optimized within the inner cross-validation loop. A multivariate feature-selection procedure identified the subset of EEG features providing the highest discriminative performance in the training data, and the optimal number of classifying features (n=50, selected in the hyperparameter selection routine, see Methods) was kept equal across standard and normative EC analyses.

**Figure 3:**
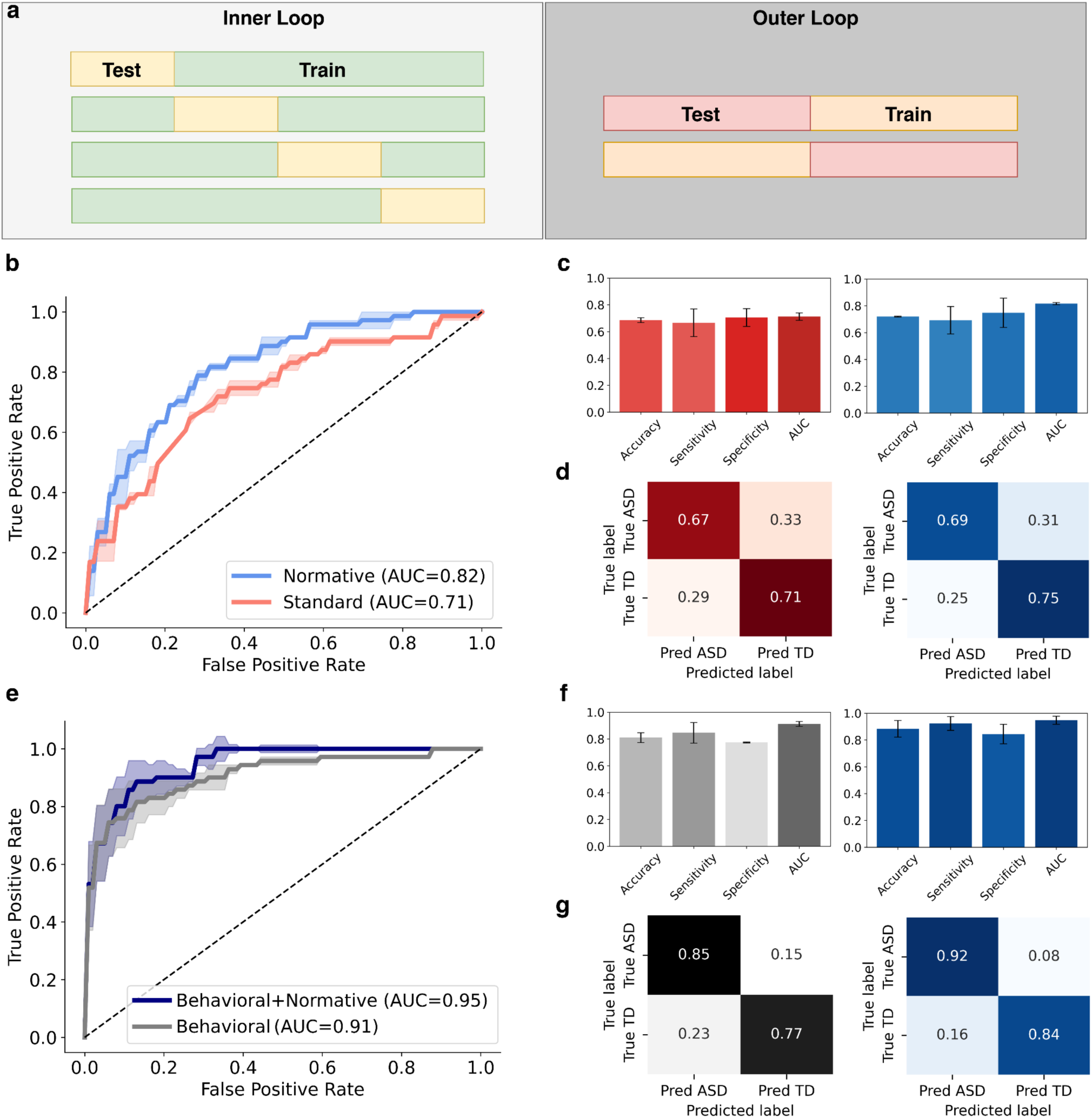
Classification (ASD vs TD) based on normative EC metrics. **(a):** Nested-cross validation architecture adopted for the machine-learning classifications. Hyperparameters and feature selection was operated in the training set of the inner loop, while performance scores were computed on the held-out test set of the outer loop. **(b):** ROC curves for the TD vs ASD classification, comparing the classifications based on standard EC and normative EC metrics (see Main text). **(c):** Performance metrics for the TD vs ASD classifications reported in (b). **(d):** Confusion matrices for the TD vs ASD classifications reported in (b). **(e):** ROC curves for the TD vs ASD classification, comparing the use of behavioral data (SRS subscales) and behavioral data combined normative EC metrics. **(f):** Performance metrics for the TD vs ASD classifications reported in (e). **(g):** Confusion matrices for the TD vs ASD classifications reported in (e). Shaded areas in (b) and (e) and error bars in (c) and (f) represent standard deviations.

On the entire dataset, classification using standard EC metrics achieved an AUC of 0.71 ± 0.03, accuracy of 0.69 ± 0.02, sensitivity of 0.71 ± 0.07, and specificity of 0.67 ± 0.10 (Figure 3b–c). Normative EC metrics improved performance, yielding an AUC of 0.82 ± 0.01, accuracy of 0.72 ± 0.01, sensitivity of 0.75 ± 0.11, and specificity of 0.69 ± 0.10 (Figure 3d). Despite this difference, no statistically significant improvement was confirmed by McNemar’s test (p = 0.125). Normative EC metrics were further evaluated using leave-one-center-out (LOCO) cross-validation, yielding AUC values of 0.71 and 0.75 on Site 1 and Site 3, respectively (mean AUC = 0.73; Site 2 was omitted as it included only TD participants).

Since normative metrics outperformed standard EC metrics in classifying ASD vs TD, we then assessed whether they also conveyed complementary information beyond standard behavioral measures. To do so, two machine-learning models (one based on SRS scores and one on SRS combined with normative EC metrics) were tested in classifying ASD vs TD participants. The model based on SRS scores alone achieved an AUC of 0.91 ± 0.02, accuracy of 0.81 ± 0.04, sensitivity of 0.78 ± 0.01, and specificity of 0.85 ± 0.08 (Figure 3e–f). Adding normative EC metrics further improved performance to an AUC of 0.95 ± 0.03, accuracy of 0.88 ± 0.06, sensitivity of 0.84 ± 0.07, and specificity of 0.92 ± 0.05 (Figure 3g). McNemar’s test confirmed a significant improvement (p = 0.004) when including normative EC metrics alongside SRS scores. LOCO cross-validation of this combined model (SRS + normative EC) yielded AUC values of 0.94 and 0.91 on Site 1 and Site 3 respectively (mean AUC = 0.93).

Notably, the combined model also supported a classification in a subset of the original dataset referring to participants younger than seven years (n=8 TD, n=8 ASD), achieving a balanced accuracy of 0.89 ± 0.04 and an AUC of 0.98, suggesting potential utility for early ASD detection.

### Normative Effective Connectivity predicts ASD symptoms severity

To investigate to which extent the normative approach could support evaluations of ASD symptoms severity, we identified a group of EC normative measures to use as multivariate predictors of SRS Total. These metrics were selected as those presenting the higher Spearman univariate correlation with SRS, and the final number of chosen measures was 8 to ensure a sufficient number of targets per each predictor (equal to n = 15 participant per predictor^37^, see Methods). Notably, the selected group was composed by measures from the same EC family (dDTF08) in beta and gamma frequency range between Right Temporo-Parietal to Right Frontal and Fronto-Frontal electrodes (Table 2).

**Table 2.** Selected EC metrics presenting high Spearman correlation with SRS metrics. Input and target channels refer to the notation Metric*_Input→Target_*.

| Metric Type | Input | Target | Frequency Band |
| --- | --- | --- | --- |
| dDTF08 | TP8 | Fp2 | Beta |
| dDTF08 | TP8 | Fp2 | Gamma |
| dDTF08 | TP8 | F4 | Beta |
| dDTF08 | TP8 | F4 | Gamma |
| dDTF08 | Fp1 | Fp2 | Beta |
| dDTF08 | Fp1 | Fp2 | Gamma |
| dDTF08 | Fp2 | Fp1 | Beta |
| dDTF08 | Fp2 | F3 | Beta |

These metrics significantly predicted SRS Total in a multilinear regression analysis (R^2^ = 0.42, p = 0.005, Figure 4a). They also predicted different SRS subscales, as confirmed by a 10-fold cross validation (Supplementary Table 3). The correlation of selected metrics with SRS subscales, however, presented no additional predictive power when the cross-correlation between SRS subscales was removed (Supplementary Table 3). To identify the EC metrics most strongly associated with symptom severity, we quantified the contribution of each selected metric to model R² using a leave-one-metric-out contribution analysis (Supplementary Table 4). The metric showing the largest contribution was right temporo-parietal (TP8) to right frontal (Fp2) beta-band effective connectivity (dDTF08_TP8→Fp2_), which explained 61.9% of the R² for SRS Total and presented the highest regression coefficient among selected metrics (Figure 4b, Supplementary Table 5, normative distribution can be visualized in Figure 2b).

**Figure 4:**
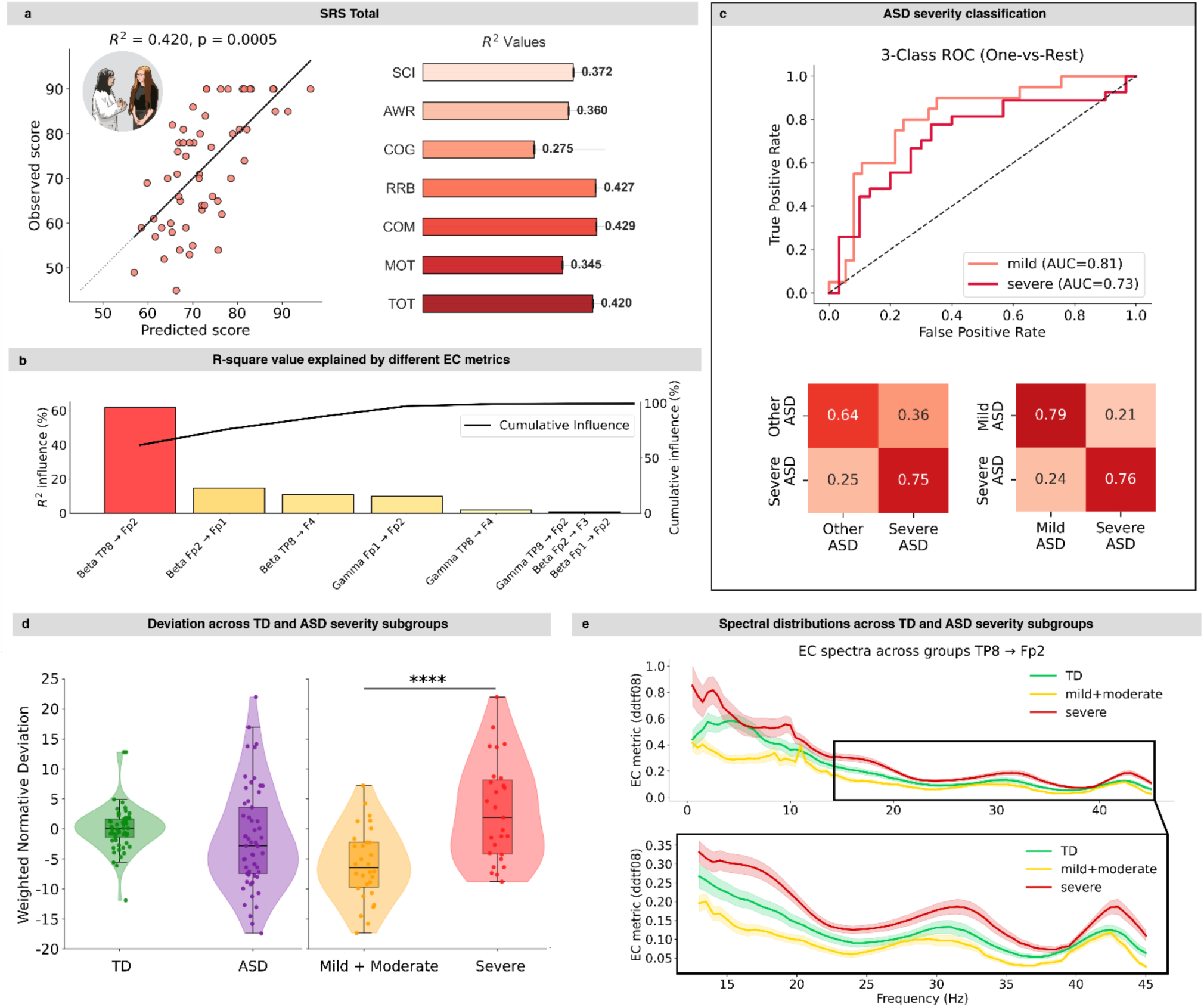
Prediction of ASD symptoms severity based on normative EC metrics. **(a):** Linear regression between selected normative EC metrics and SRS Total values. (left) with cumulative R² values for different SRS subscales reported on the right. Reported R² values are not cross-validated (cross-validated versions are reported in Supplementary Table 3). **(b):** Pareto chart for the influence of each of the selected metrics with respect to the R^2^ of SRS Total scale in ASD subjects. All EC metrics pertain to the DTF family, most sensitive to neural signal sinks (i.e. electrodes receiving high total inputs). The Pareto of each subscale can be found in Supplementary Figure 2. **(c):** ROC curves and confusion matrices for (i) the classification between severe ASD and other ASD (mild and moderate), and for (ii) the classification between severe ASD and mild ASD **(d):** Left: Boxplot distribution of the Weighted Normative Deviation for TD (green) and ASD (purple). Right: Boxplot distribution of the WND for mild & moderate ASD (yellow) and severe ASD (red). Each point of the box-plot represents a participant. Horizontal lines represent median values, while whiskers are set at 1.5 times the interquartile range. Distributions are over-imposed and were calculated with kde. **(e):** Frequency spectrum of the main contributor to SRS scales (Beta-band dDTF08 _TP8->Fp2_) divided by group severity, TD (green), mild & moderate ASD (yellow), severe ASD (red); bold lines represent average, shaded areas representing the Standard Error of the Mean (SEM); in beta and gamma bands (13-45 Hz) the clinical groups are quite well ordered respect to TD.

Since this metric alone contributed more than the others combined in determining the total R^2^ value, we tested it to differentiate between different ASD severity subgroups. Using this metric as a single classifying feature with a moving threshold, severe ASD participants (SRS Total > 75, n=30) were classified against mild/moderate ASD participants (SRS Total ≤ 75, n=27, Figure 4c), yielding an AUC of 0.73, sensitivity of 0.75, specificity of 0.64, and balanced accuracy of 0.69. Restricting the comparison to severe ASD versus mild ASD (SRS Total T<65, n=20) individuals further improved performance (AUC=0.81, sensitivity=0.76, specificity=0.79, balanced accuracy=0.78). This metric thus consistently differentiated between ASD severity subgroups and predicted different SRS subscales with values higher than those of other selected metrics (Figure 4c, Supplementary Figure 2).

We then examined raw EC values prior to normative modeling in the initial groups of 64 TD and 64 ASD participants across the full frequency spectrum (0.5–45 Hz), grouping subjects by clinical severity (TD; mild/moderate ASD, n=27; severe ASD, n=30). The Beta-band dDTF08 TP8→Fp2 metric showed a clear severity-dependent ordering in the beta and gamma range (13–45 Hz): severe ASD exhibited relative hyperconnectivity compared with TD, while mild/moderate ASD showed relative hypoconnectivity (Figure 4e). This pattern likely underlies the monotonic association between EC measures and clinical severity within the ASD group, as supported by significant Spearman and Pearson correlations in the predicted-versus-observed SRS analyses (SRS Total: r=0.65, p<0.00001; r=0.66, p<0.00001).

We next examined the distribution of weighted normative deviation scores in TD and ASD groups using group-specific regression weights from the SRS Total models (Figure 4d, left). No statistically significant differences were found between groups, although ASD participants presented a broader distribution (median_TD_ = −0.11, MAD_TD_ = 1.57; median_ASD_ = −2.83, MAD_ASD_ = 4.96). A further comparison of intra-group variabilities made using the Levene’s test corroborated this result, showing a significantly greater heterogeneity in the ASD group (F = 26.2, p < 0.00001). Stratifying the ASD group into mild/moderate and severe subgroups revealed significantly higher weighted deviation scores in the severe subgroup (Mann–Whitney U = 161, p = 0.0001, Cohen’s d = 1.22; Figure 4d, right), with the largest differences concentrated in the beta and gamma bands (Figure 4e).

A complementary principal component analysis (PCA) exploring the relationship between normative EC metrics and SRS subscales, compared with results from standard EC metrics, is reported in Supplementary Materials (Supplementary Figure 3). Symptom-specific association of normative EC metrics was also performed. Results are reported in Supplementary Materials (Supplementary Figures 4-5). Briefly, normative EC metrics predicted different SRS subscales significantly outperforming standard EC metrics in both groups (paired t-test: t=-4.63, p=0.002, d=1.54 for ASD; t=-4.32, p=0.003, d=1.44 for TD), also presenting significant topographic association with distinct SRS subscales.

## Discussion

Our results support the use of normative EC metrics as robust ASD neural markers, enabling both accurate classification of ASD participants and robust, symptom-specific associations with behavioral subscales.

A core aspect of our study is the identification of robust, symptom-specific EEG normative EC metrics capable of predicting SRS subscales with high precision (R^2^=0.42 for SRS Total). Other studies investigated the prediction of behavioral metrics such as SRS using quantitative neural markers. Zhang et al.^38^ predicted Autism Diagnostic Observation Schedule (ADOS) values using a linear regression model based on EEG functional connectivity in 257 ASD participants, with r-values ranging from 0.20 to 0.40. ADOS repetitive behaviors scores were also predicted using EEG functional connectivity metrics by Tong et al.^39^ in 392 ASD participants, with an r-value of 0.45. Longitudinal ADOS changes were also predicted in 188 TD participants (including 99 participants with ASD familiarity) using quantitative EEG metrics by Bosl et al.^40^. Pereira et al.^41^ reported a prediction of Autism Diagnostic Interview-Revised (ADI-R) scores based on fMRI quantities with r-values ranging from 0.50 to 0.63 in 22 ASD participants.

Our study highlighted how EEG normative EC metrics can support a robust ASD vs TD classification, both used on their own and in combination with behavioral scales. The classification between ASD and TD participants using neural makers has been vastly investigated in recent literature. Comparing our results with a meta-analysis based on fMRI studies^42^, our ASD vs TD classification results based on SRS combined with EEG normative EC metrics exceeded the upper 95% confidence interval in ASD diagnosis based on combining fMRI and behavioral metrics (balanced accuracy: 0.878 vs. our results of balanced accuracy = 0.88).

Our analysis identified a subset of normative EC metrics predicting behavioral scales with high significance (Figure 4a). These selected metrics form a coherent and biologically plausible group of neural markers pertaining to an EC family (dDTF08) highly sensitive to connectivity sinks, i.e. brain regions (or electrodes) which receive great magnitude of input from surrounding areas. Alterations in DTF metrics suggest anomalies in integration and segregation of neural information, as confirmed by recent ASD research^43,44^. These selected metrics were all related to Beta- and Gamma-band, which have been widely explored in the literature, particularly for the association of attentional and sensorimotor processes for the Beta-band^45^ and of behavioral scales (including SRS) for the Gamma-band^46^.

Importantly, these connectivity patterns are also supported by prior literature, helping to interpret their neurobiological meaning (Barnes et al.^26^; Van Overwalle^47^). Also Van Overwalle^47^ highlighted the central role of functional connectivity involving the medial prefrontal cortex (mPFC), temporoparietal junction, and posterior superior temporal sulcus, together with subcortical structures, in social and emotional learning, two core domains strongly affected in ASD. This framework guided the selection of the six electrodes used in the present study, as they showed the closest anatomical correspondence to these regions, see Supplementary Figure 6. A similarity analysis between our findings and those of Barnes et al.^26^ is reported in Supplementary Materials.

The opposite trend with respect to TD observed in severe and mild/moderate ASD (Figure 4d) provides additional support for the hypothesis that ASD may be better conceptualized as a high-variance distribution in which each individual deviates from the neurotypical developmental trajectory in distinct ways. In this framework, different combinations of disrupted neural mechanisms may converge onto apparently similar scores on the same behavioral phenotypes, while still preserving markedly greater dispersion relative to TD. This interpretation is consistent with the idiosyncrasy framework recently proposed by Lin et al.^33^

Several limitations of the present study should be acknowledged. First, the normative modeling approach was derived from a relatively small sample of TD participants, and normative modelling and classification procedures were not fully nested within the same cross-validation framework. This may limit the robustness and generalizability of the estimated normative distributions. Future studies should validate these findings using different cohorts for normative modelling and classification to prevent data leakage. Second, all data were drawn from the same large-scale dataset collected in the United States, limiting sociocultural and environmental diversity. Validation on independent datasets from different contexts will therefore be essential. Third, the analysis of EC metrics depends on the selection of EEG channels: although channels were chosen based on prior literature and known pathophysiological mechanisms in ASD, alternative configurations could yield different connectivity patterns and associations with behavioral measures. Finally, the study focused on participants aged 5–19 years, excluding younger age ranges that are critical for early ASD development and diagnosis. Extending this approach to younger populations will be necessary to assess the potential utility of these metrics for early identification.

## Conclusions

EEG Normative EC metrics emerged as robust ASD neural markers, capable of both supporting a classification of ASD and TD participants and relating to the severity of symptoms. The combination with SRS scores in ASD classification allowed to obtain high inter-center accuracy, outperforming the accuracy of behavioral scales alone. EEG Normative EC metrics could potentially provide key low-cost and interpretable neural markers, supporting the diagnosis and assessment of ASD severity.

## Methods

### Participants

All participants included in this study were drawn from the Healthy Brain Network (HBN) dataset^35^, for a total of 149 subjects. Of these, 9 subjects did not pass the preprocessing phase and were excluded from subsequent analyses.

We selected 64 ASD and 64TD to obtain a demographically controlled case-control subset^28^, matched for sex and stratified by age, in order to minimize confounding and support both between-group statistical comparisons and clinical regression. In the initial matched cohort, matching quality was verified by confirming no significant between-group differences in age (Welch’s t-test, p = 0.89) and sex distribution (χ² test, p = 1.00). Balance across the predefined age strata was additionally confirmed by negligible standardized mean differences across strata and perfect sex balance within strata (|SMD_AGE_| ≤ 0.22; |SMD_SEX_| = 0.00 across strata).

This sample size was consistent with a priori power considerations for detecting at least large group effects (α = 0.05 before multiple-comparison correction, power = 0.80, effect size d = 0.80, allocation ratio N2/N1 = 1, using G*Power3.1). In the sample size estimation, we additionally accounted for multiple-comparison control by adopting a conservative Bonferroni-corrected significance threshold (N = 30 tests), although the final statistical analyses were controlled using the Benjamini–Hochberg false discovery rate (FDR-BH) procedure.

After preprocessing and quality-control steps, statistical analyses were restricted to subjects with complete EC and SRS data, yielding the final analytical subset of 57 ASD and 68 TD participants. To ensure a fair comparison between, groups, a final subset of 57 TD subjects was identified to minimize sex and age differences across groups. In this final analytical subset, matching quality remained acceptable, with no significant between-group differences in age (Welch’s t-test, p = 0.68) or sex distribution (χ² test, p = 0.47). Balance across the predefined age strata remained acceptable, with small standardized mean differences across all strata (|SMD_AGE_| ≤ 0.22 and |SMD_SEX_| ≤ 0.33); age and sex were additionally included as covariates in the statistical models. All data can be found in Supplementary Table 6.

It should also be noted that, during the ASD–TD group balancing procedure for age and sex, 12 TD subjects were excluded due to group imbalance. However, given the limited number of TD subjects available for constructing the normative reference curves, these subjects were retained for normative modelling purposes only and were not included in subsequent group comparisons or clinical regression analyses.

### EEG recordings and preprocessing

Resting-state EEG was acquired from the Healthy Brain Network (HBN) dataset using a 128-channel HydroCel Geodesic Sensor Net (Electrical Geodesics Inc., Eugene, OR, USA) with a sampling frequency of 500 Hz. For each subject, the eyes-closed resting-state intervals were identified from the native event structure within the alternating eyes-open/eyes-closed protocol. Preprocessing was performed in MATLAB (MathWorks, Natick, MA, USA) using EEGLAB with the plugins clean_rawdata, ICLabel, and SIFT. Signals were first high-pass filtered at 0.5 Hz and low-pass filtered at 45 Hz using zero-phase FIR Hamming filters (pop_eegfiltnew), followed by a notch filter (58–62 Hz) to attenuate line noise. Automated artifact rejection was then applied using clean_rawdata, including removal of flat or poorly correlated channels, Artifact Subspace Reconstruction (ASR), and contaminated windows. Data were subsequently re-referenced to the common average reference. Independent Component Analysis (ICA) was performed using the FastICA algorithm, and independent components were automatically classified with ICLabel. Given the high channel density of the recordings, objective pre-marking rules were applied to reduce the burden of manual inspection. Components with Brain probability <50%, components assigned to non-brain classes (Eye, Muscle, Heart, Line Noise, and Channel Noise), and components classified as “Other” with classification confidence <75% were pre-marked for rejection. A subsequent visual inspection of component features such as PSD and brain functional topography was then performed before final component removal. Channel labels were standardized using the adjusted HydroCel montage, and peripheral/non-cortical electrodes were removed to retain only cortical scalp channels for subsequent analyses. For each of the five eyes-closed periods, the first 5 s were discarded to avoid transition effects, and the subsequent 10 s were extracted, yielding five candidate 10-s resting-state segments per subject. These segments were used to compute empirical EEG features in Python (MNE and MNE-connectivity), including power spectral density (PSD) and functional connectivity (Pearson correlation). The representative segment was defined as the 10-s epoch minimizing a subject-specific loss function computed as the average distance between its PSD and FC profiles and the corresponding mean PSD and FC profiles across all available segments of the same subject.

For the subsequent effective connectivity estimation, the limited amount of available data required the selection of a reduced subset of channels among the 128 electrodes available in the HBN dataset. This is because multivariate autoregressive (MVAR) modeling requires a sufficient number of time points (tp) relative to the number of modeled channels (p) and the selected model order (n) to ensure stable parameter estimation (ideally tp >> n x p^2^). Accordingly, we adopted a hypothesis-driven approach and selected only six electrodes (TP7, TP8, F3, F4, Fp1, and Fp2) based on the available literature (see Discussion for the rationale underlying this selection).

### Effective Connectivity Estimation

Effective connectivity was estimated in a subset of six channels (TP7, TP8, F3, F4, Fp1, Fp2) using a multivariate autoregressive Granger Causality (MVAR GC) EEG modeling^48^ approach implemented in the SIFT toolbox^36^ of EEGLAB. For each EEG dataset a resting state eyes closed window of circa 10s was used; an MVAR model was fitted using the ARfit algorithm, which is well-suited for multichannel electrophysiological data.

The model order was initially explored within a broad range (1-50) using multiple information criteria^36^ (AIC, SBC/BIC, FPE, and HQ). However, automatic selection often resulted in relatively low model orders that failed to adequately capture the temporal structure of the data, as indicated by poor signal reconstruction consistency and stability. Therefore, a fixed model order of 20 was adopted for all subjects. This choice ensured high reconstruction accuracy (approximately 95% consistency between empirical and modeled signals) and improves the stability of model validation metrics with a trade-off for residual whiteness. Using a fixed order also enhances comparability across subjects, avoiding variability introduced by subject-specific model order estimation.

Model validation was systematically performed to ensure the reliability of the estimated parameters. Specifically, residual whiteness was assessed using multiple complementary tests^36^ (Ljung-Box, Box-Pierce, autocorrelation function inspection, and Li-McLeod multivariate test), confirming that the MVAR models adequately captured the temporal dependencies of the data. In addition, model stability was verified by ensuring that all eigenvalues of the system lay within the unit circle, and the percentage of data consistency (i.e., variance explained by the model) was computed as an additional goodness-of-fit measure.

From the validated MVAR models, the effective connectivity measures^36^ reported in Table 1 were calculated. These metrics were computed across a frequency range of interest using the estimated model coefficients (0.5 – 45 Hz). To mitigate the impact of volume conduction and spurious zero-lag correlations, a phase random permutation (random rephasing) procedure was applied to generate surrogate data, allowing for statistical thresholding of connectivity estimates (500 permutations, alpha=0.05 corrected for multiple comparisons using FDR Benjamini Hochberg, more details can be found in the corresponding SIFT publication^36^ and website: https://eeglab.org/plugins/SIFT/ ).

Finally, connectivity matrices were obtained for each subject and metric, and subsequent analyses were performed on these matrices after correction for multiple comparisons (FDR BH). All steps were applied consistently across subjects to ensure methodological robustness and reproducibility. Robustness analysis regarding the dependence of EC metric values on the model order parameter, and their inter-subject variability is reported in Supplementary Materials.

### Normative modeling of effective connectivity

To quantify subject-specific deviations in effective connectivity relative to typical neurodevelopmental trajectories, a normative modeling framework based on Gaussian Process Regression (GPR)^49,50^ was applied with a radial basis function (RBF, squared exponential) kernel and additive white-noise term. The model was trained exclusively on typically developing (TD) participants and subsequently used to estimate individualized deviations in both TD and autism spectrum disorder (ASD) subjects.

Age was modeled as a continuous variable and used as the only covariate in the normative model, given its major influence on developmental EEG connectivity patterns and to preserve model stability considering the available TD sample size. Sex was not included as an explicit predictor in the primary model to avoid overparameterization and imbalance effects within the control cohort.

For each EC feature independently, a GPR model was fitted to estimate the expected age-dependent normative trajectory and its associated predictive uncertainty. In TD participants, out-of-sample normative estimates were obtained using 5-fold cross-validated predictions (zscorecv), ensuring unbiased estimation of normative deviations. For ASD participants, the final normative model trained on the full TD cohort was used to compute subject-level deviations relative to the reference population.

Individual deviation scores were expressed as standardized z-scores:

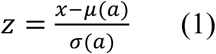

where x represents the observed EC value, m(a) the age-specific normative expectation estimated by the GPR model, and s(a) the corresponding predictive standard deviation. For TD participants, m(a) and s(a) were obtained from out-of-sample 5-fold cross-validated predictions, whereas for ASD participants they were obtained from the model of the full TD cohort. Positive and negative z-scores indicate hyperconnectivity and hypoconnectivity, respectively, relative to the normative developmental trajectory. Normative curves of the selected metrics reported in the *“Normative Effective Connectivity predicts ASD symptoms severity”* can be found in Supplementary Figure 7. The group of EC normative measures was selected based on the Spearman univariate correlation with SRS, leading to the group of Direct Transfer Function metrics reported in Table 2.

These subject-specific normative deviation scores were subsequently used for clinical association analyses with SRS-2 total and subscale T-scores, subgroup stratification, and computation of the weighted normative deviation (WND), a composite subject-level biomarker integrating multimetric EC abnormalities according to their clinical regression coefficients.

### Machine-Learning classifications

Machine learning classification analyses were performed to discriminate between groups based on features derived from EEG data and model-based parameters. A logistic regression was adopted as the primary classifier, given its robustness in settings characterized by relatively small sample sizes and high-dimensional feature spaces, as well as its interpretability in terms of linear weights. Hyperparameters chosen for the classification (which we report here in brackets) were the penalty (“l2”), the solver (“sag”), and the number of max iterations (2000), alongside the total number of classifying features (50), while the regularization parameter “C” was selected automatically (see below). Hyperparameters were kept equal for all classifications and were selected in the TD vs. ASD classification (except for the number of features for the TD vs. ASD classification based on behavioral metrics, which was limited by the total number of SRS subscales plus the age of patients). Feature selection was operated in the training set of the inner loop (Figure 3A), with the total number of classifying features selected automatically (see below). When appropriate, features were normalized to a [0, 1] range to ensure comparability across different scales.

Hyperparameter tuning, specifically for the regularization parameter C of the Logistic Regression and for the total number of classifying features, was carried out using a nested cross-validation framework to prevent optimistic bias in performance estimation. While the number of classifying features was kept equal across all classifications, the C parameter was selected autonomously at each loop to find the optimal value. The outer loop consisted of a 2-fold cross-validation scheme used to assess the generalization performance of the model (Figure 3a). Within each training fold, an inner 4-fold cross-validation loop was implemented to optimize hyperparameters via grid search in the training set.

Model performance was quantified in the test set of the outer loop using multiple metrics, including accuracy, AUC score, sensitivity, and specificity. Given the potential class imbalance in the dataset, particular emphasis was placed on AUC and balanced accuracy. To assess the statistical significance of the observed performance, the cross-validation procedure was repeated multiple times with different random splits. The resulting distributions of performance metrics were then compared across models or feature sets using non-parametric statistical tests (e.g., Mann-Whitney U test). When multiple comparisons were performed, p-values were corrected using the FDR procedure.

To further evaluate the robustness and generalizability of the classifier across different acquisition centers, a LOCO cross-validation strategy was implemented. In this approach, data from one clinical center were iteratively held out as an independent test set, while the model was trained on data from the two remaining centers. This procedure provided a stringent assessment of cross-site generalization and better reflects real-world clinical deployment scenarios, where models are required to generalize to unseen data acquired under different conditions.

### Statistical Analysis

All statistical analyses were performed using standard Python libraries (Numpy, SciPy, StatsModels). Demographic and clinical characteristics of participants were compared between ASD and TD groups using group-level statistical analyses on the final analytical subset with complete EC and SRS data. Continuous variables, including age and SRS-2 T-scores, were assessed using two-sided non-parametric Mann–Whitney U tests when appropriate, while age matching in the subset was additionally verified using Welch’s t-test. Categorical variables, including sex distribution, were evaluated using chi-square tests. Matching quality across predefined age strata was further quantified using standardized mean differences (SMD) for age and sex. Group differences between ASD and TD participants were assessed using non-parametric Mann–Whitney U tests for continuous variables, due to potential deviations from normality, and chi-square tests for categorical variables. Effect sizes were quantified using Cohen’s d for continuous measures and appropriate association metrics for categorical variables. Differences in intra-group variabilities were assessed using Levene’s test. To account for multiple comparisons across features, all p-values were corrected using the false discovery rate (FDR) procedure according to the Benjamini–Hochberg method.

To account for potential confounding effects, additional analyses incorporating covariates were conducted using generalized linear models. Specifically, age, sex, and acquisition site were included as covariates, and interaction terms (e.g., Group × Site) were explicitly modeled to assess whether group differences varied across centers. Statistical significance of interaction effects was evaluated based on the corresponding p-values of the interaction terms within the model. Statistical differences between classifications were based on the McNemar’s test.

Normative metrics employed as multivariate SRS predictors were selected based on their Spearman correlation with the target variable. Their performance was assessed using R² between observed and predicted values. To evaluate generalization performance and reduce overfitting, a total number of 8 multivariate predictors was used to ensure at least a 1/15 ratio between predictors and targets^37^. A 10- fold cross-validation was applied. Whenever possible, folds were stratified according to binned SRS distributions to preserve balanced symptom severity across folds. Out-of-fold (OOF) predictions were used to compute cross-validated performance metrics, including R², Pearson’s r, and Spearman’s ρ. Only complete-case subjects with available EC and SRS data were included in the regression analyses.

Dimensionality reduction (Supplementary Figure 3) was performed using PCA on selected feature subsets to identify dominant patterns of variance. Linear regression models were then applied to evaluate the relationship between principal components and clinical or group variables. These models included the same covariates (age, sex, and site) to control for potential confounds. Regression coefficients, associated p-values, and model fit statistics were reported, with FDR correction applied across multiple components. Linear regression analyses were also conducted with single normative EC metrics to evaluate the relationship with SRS-2 Total and subscales T-scores. Ordinary least squares (OLS) models were fitted using normative EC features as predictors, with optional inclusion of age and sex as covariates depending on the specific model.

Feature-specific contributions to symptom prediction were quantified using a leave-one-metric-out ΔR² analysis: for each normative EC feature, its contribution was estimated as the reduction in model R² after removing that feature from the full regression model predicting SRS Total T-score. Relative contributions were normalized and visualized using Pareto plots with cumulative explained variance. Weighted normative deviation scores were computed as the dot product between the multifeature normative deviation vector and the regression coefficients derived from the SRS Total T-score model, using group-specific coefficients for TD and ASD subjects. Median values and median absolute deviations (MAD) were additionally reported for descriptive purposes.

Finally, symptom-specific associations were explored by correlating normative EC metrics with SRS subscales, considering only metrics with an R² threshold of 0.42, which was determined from the value for SRS Total prediction obtained with selected EC metrics. For each subscale, multiple comparisons across connectivity features were controlled using FDR correction. Additionally, partial correlation was employed to adjust for the previously mentioned covariates. This analysis allowed for the identification of connectivity patterns selectively associated with specific symptom domains, providing a more fine-grained characterization of the relationship between brain connectivity alterations and behavioral phenotypes in ASD. A robustness analysis of the computed EC metrics was also performed. Results are reported in Supplementary Materials (Supplementary Figure 8).

## Data Availability

All data produced in the present study are available upon reasonable request to the authors.
All HBN data are available at https://fcon_1000.projects.nitrc.org/indi/cmi_healthy_brain_network/

https://fcon_1000.projects.nitrc.org/indi/cmi_healthy_brain_network/

## Ethical approval

In our study, we analyzed EEG and phenotypic data from data releases 1–11 of the Healthy Brain Network (HBN) biobank^35^. HBN is an initiative of the Child Mind Institute to establish a large-scale biobank of individuals aged 5–21 years recruited from the New York metropolitan area. Further details on the HBN biobank, including experimental procedures and scan parameters, are provided in ^35^. The HBN study was approved by the Chesapeake Institutional Review Board, and written informed consent was obtained from all participants and/or their legal guardians.

## References

1. Lord C, Brugha TS, Charman T, et al. Autism spectrum disorder. Nat Rev Dis Primers. 2020;6(1):5. doi:10.1038/s41572-019-0138-4

2. Lord C, Elsabbagh M, Baird G, Veenstra-Vanderweele J. Autism spectrum disorder. The Lancet. 2018;392(10146):508-520. doi:10.1016/S0140-6736(18)31129-2

3. Santomauro DF, Erskine HE, Mantilla Herrera AM, et al. The global epidemiology and health burden of the autism spectrum: findings from the Global Burden of Disease Study 2021. The Lancet Psychiatry. 2025;12(2):111–121. doi:10.1016/S2215-0366(24)00363-8

4. Zeidan J, Fombonne E, Scorah J, et al. Global prevalence of autism: A systematic review update. Autism Res. 2022;15(5):778–790. doi:10.1002/aur.2696

5. Shaw KA. Prevalence and Early Identification of Autism Spectrum Disorder Among Children Aged 4 and 8 Years — Autism and Developmental Disabilities Monitoring Network, 16 Sites, United States, 2022. MMWR Surveill Summ. 2025;74. doi:10.15585/mmwr.ss7402a1

6. Hus Y, Segal O. Challenges Surrounding the Diagnosis of Autism in Children. Neuropsychiatric Disease and Treatment. 2021;17:3509–3529. doi:10.2147/NDT.S282569

7. Barbaro J, Halder S. Early Identification of Autism Spectrum Disorder: Current Challenges and Future Global Directions. Curr Dev Disord Rep. 2016;3(1):67–74. doi:10.1007/s40474-016-0078-6

8. Frye RE, Vassall S, Kaur G, Lewis C, Karim M, Rossignol D. Emerging biomarkers in autism spectrum disorder: a systematic review. Ann Transl Med. 2019;7(23):792. doi:10.21037/atm.2019.11.53

9. Oxelgren UW, Myrelid Å, Annerén G, et al. Prevalence of autism and attention-deficit-hyperactivity disorder in Down syndrome: a population-based study. Dev Med Child Neurol. 2017;59(3):276–283. doi:10.1111/dmcn.13217

10. Warner G, Moss J, Smith P, Howlin P. Autism characteristics and behavioural disturbances in ∼ 500 children with Down’s syndrome in England and Wales. Autism Res. 2014;7(4):433–441. doi:10.1002/aur.1371

11. Traut N, Heuer K, Lemaître G, et al. Insights from an autism imaging biomarker challenge: Promises and threats to biomarker discovery. NeuroImage. 2022;255:119171. doi:10.1016/j.neuroimage.2022.119171

12. Emerson RW, Adams C, Nishino T, et al. Functional neuroimaging of high-risk 6-month-old infants predicts a diagnosis of autism at 24 months of age. Sci Transl Med. 2017;9(393):eaag2882. doi:10.1126/scitranslmed.aag2882

13. Hazlett HC, Gu H, Munsell BC, et al. Early brain development in infants at high risk for autism spectrum disorder. Nature. 2017;542(7641):348–351. doi:10.1038/nature21369

14. James SJ, Melnyk S, Jernigan S, et al. Metabolic endophenotype and related genotypes are associated with oxidative stress in children with autism. Am J Med Genet B Neuropsychiatr Genet. 2006;141B(8):947-956. doi:10.1002/ajmg.b.30366

15. 15. Bogéa Ribeiro L, da Silva Filho M. Systematic Review on EEG Analysis to Diagnose and Treat Autism by Evaluating Functional Connectivity and Spectral Power. Neuropsychiatr Dis Treat. 2023;19:415-424. doi:10.2147/NDT.S394363

16. Lyall K. What are quantitative traits and how can they be used in autism research? Autism Research. 2023;16(7):1289–1298. doi:10.1002/aur.2937

17. Hirota T, King BH. Autism Spectrum Disorder: A Review. JAMA. 2023;329(2):157–168. doi:10.1001/jama.2022.23661

18. Constantino JN. Social Responsiveness Scale. In: Encyclopedia of Autism Spectrum Disorders. Springer, Cham; 2021:4457–4467. doi:10.1007/978-3-319-91280-6_296

19. Bölte S, Poustka F, Constantino JN. Assessing autistic traits: cross-cultural validation of the social responsiveness scale (SRS). Autism Research. 2008;1(6):354–363. doi:10.1002/aur.49

20. Hus V, Bishop S, Gotham K, Huerta M, Lord C. Factors influencing scores on the social responsiveness scale. Journal of Child Psychology and Psychiatry. 2013;54(2):216–224. doi:10.1111/j.1469-7610.2012.02589.x

21. Wang J, Barstein J, Ethridge LE, Mosconi MW, Takarae Y, Sweeney JA. Resting state EEG abnormalities in autism spectrum disorders. J Neurodevelop Disord. 2013;5(1):24. doi:10.1186/1866-1955-5-24

22. Coben R, Clarke AR, Hudspeth W, Barry RJ. EEG power and coherence in autistic spectrum disorder. Clinical Neurophysiology. 2008;119(5):1002–1009. doi:10.1016/j.clinph.2008.01.013

23. Bosl W, Tierney A, Tager-Flusberg H, Nelson C. EEG complexity as a biomarker for autism spectrum disorder risk. BMC Med. 2011;9(1):18. doi:10.1186/1741-7015-9-18

24. Righi G, Tierney AL, Tager-Flusberg H, Nelson CA. Functional Connectivity in the First Year of Life in Infants at Risk for Autism Spectrum Disorder: An EEG Study. PLOS ONE. 2014;9(8):e105176. doi:10.1371/journal.pone.0105176

25. Salehi F, Jaloli M, Coben R, Nasrabadi AM. Estimating brain effective connectivity from EEG signals of patients with autism disorder and healthy individuals by reducing volume conduction effect. Cogn Neurodyn. 2022;16(3):519–529. doi:10.1007/s11571-021-09730-w

26. Barnes SJK, Thomas M, McClintock PVE, Stefanovska A. Theta and alpha connectivity in children with autism spectrum disorder. Brain Commun. 2025;7(2):fcaf084. doi:10.1093/braincomms/fcaf084

27. Rutherford S, Barkema P, Tso IF, et al. Evidence for embracing normative modeling. Baker CI, Constable T, Esteban O, eds. eLife. 2023;12:e85082. doi:10.7554/eLife.85082

28. Marquand AF, Kia SM, Zabihi M, Wolfers T, Buitelaar JK, Beckmann CF. Conceptualizing mental disorders as deviations from normative functioning. Mol Psychiatry. 2019;24(10):1415–1424. doi:10.1038/s41380-019-0441-1

29. Mandelli V, Landi I, Busuoli EM, Courchesne E, Pierce K, Lombardo MV. Prognostic early snapshot stratification of autism based on adaptive functioning. Nat Mental Health. 2023;1(5):327–336. doi:10.1038/s44220-023-00056-6

30. Zabihi M, Floris DL, Kia SM, et al. Fractionating autism based on neuroanatomical normative modeling. Transl Psychiatry. 2020;10(1):384. doi:10.1038/s41398-020-01057-0

31. Bethlehem RAI, Seidlitz J, Romero-Garcia R, Trakoshis S, Dumas G, Lombardo MV. A normative modelling approach reveals age-atypical cortical thickness in a subgroup of males with autism spectrum disorder. Commun Biol. 2020;3(1):486. doi:10.1038/s42003-020-01212-9

32. Lombardo MV, Severino I, Mandelli V. Stratifying the autisms by a type I versus type II distinction in early development. Nat Mental Health. 2026;4(3):321–335. doi:10.1038/s44220-026-00603-x

33. Lin HY, Breakspear M, Mottron L. From heterogeneity to idiosyncrasy in the autistic brain. Nat Mental Health. 2026;4(3):346–359. doi:10.1038/s44220-026-00601-z

34. Jiang A, Ma X, Li S, et al. Age-atypical brain functional networks in autism spectrum disorder: a normative modeling approach. Psychological Medicine. 2024;54(9):2042–2053. doi:10.1017/S0033291724000138

35. Alexander LM, Escalera J, Ai L, et al. An open resource for transdiagnostic research in pediatric mental health and learning disorders. Sci Data. 2017;4(1):170181. doi:10.1038/sdata.2017.181

36. Delorme A, Mullen T, Kothe C, et al. EEGLAB, SIFT, NFT, BCILAB, and ERICA: new tools for advanced EEG processing. Comput Intell Neurosci. 2011;2011:130714. doi:10.1155/2011/130714

37. Riley RD, Snell KI, Ensor J, et al. Minimum sample size for developing a multivariable prediction model: PART II - binary and time-to-event outcomes. Statistics in Medicine. 2019;38(7):1276–1296. doi:10.1002/sim.7992

38. Zhang Y, Zhang S, Chen B, et al. Predicting the Symptom Severity in Autism Spectrum Disorder Based on EEG Metrics. IEEE Transactions on Neural Systems and Rehabilitation Engineering. 2022;30:1898–1907. doi:10.1109/TNSRE.2022.3188564

39. Tong X, Xie H, Fonzo GA, et al. Symptom dimensions of resting-state electroencephalographic functional connectivity in autism. Nat Mental Health. 2024;2(3):287–298. doi:10.1038/s44220-023-00195-w

40. Bosl WJ, Tager-Flusberg H, Nelson CA. EEG Analytics for Early Detection of Autism Spectrum Disorder: A data-driven approach. Sci Rep. 2018;8(1):6828. doi:10.1038/s41598-018-24318-x

41. Pereira AM, Campos BM, Coan AC, et al. Differences in Cortical Structure and Functional MRI Connectivity in High Functioning Autism. Front Neurol. 2018;9. doi:10.3389/fneur.2018.00539

42. Santana CP, de Carvalho EA, Rodrigues ID, Bastos GS, de Souza AD, de Brito LL. rs-fMRI and machine learning for ASD diagnosis: a systematic review and meta-analysis. Sci Rep. 2022;12(1):6030. doi:10.1038/s41598-022-09821-6

43. Yang B, Wang M, Zhou W, et al. Disrupted network integration and segregation involving the default mode network in autism spectrum disorder. Journal of Affective Disorders. 2023;323:309–319. doi:10.1016/j.jad.2022.11.083

44. Rudie JD, Shehzad Z, Hernandez LM, et al. Reduced Functional Integration and Segregation of Distributed Neural Systems Underlying Social and Emotional Information Processing in Autism Spectrum Disorders. Cereb Cortex. 2012;22(5):1025–1037. doi:10.1093/cercor/bhr171

45. Neo WS, Foti D, Keehn B, Kelleher B. Resting-state EEG power differences in autism spectrum disorder: a systematic review and meta-analysis. Transl Psychiatry. 2023;13(1):389. doi:10.1038/s41398-023-02681-2

46. Rojas DC, Wilson LB. γ-Band Abnormalities as Markers of Autism Spectrum Disorders. Biomarkers in Medicine. 2014;8(3):353–368. doi:10.2217/bmm.14.15

47. Van Overwalle F. Social and emotional learning in the cerebellum. Nat Rev Neurosci. 2024;25(12):776–791. doi:10.1038/s41583-024-00871-5

48. Anderson CW, Stolz EA, Shamsunder S. Multivariate autoregressive models for classification of spontaneous electroencephalographic signals during mental tasks. IEEE Transactions on Biomedical Engineering. 1998;45(3):277–286. doi:10.1109/10.661153

49. Ebadi A, Allouch S, Mheich A, et al. Beyond homogeneity: charting the landscape of heterogeneity in neurodevelopmental and psychiatric electroencephalography. Transl Psychiatry. 2025;15(1):223. doi:10.1038/s41398-025-03441-0

50. Fraza C, Rutherford S, Bučková BR, Beckmann CF, Marquand AF. The promise of quantifying individual risk for brain disorders through normative modeling, a narrative review. Neuroscience & Biobehavioral Reviews. 2025;176:106284. doi:10.1016/j.neubiorev.2025.106284

