## Supplementary_Material for "Normative EEG Effective Connectivity as a neural marker of Autism Spectrum Disorder"

### Supplementary Materials:

#### Supplementary Results

##### Normative EC PCA-based Analysis of SRS subscales

To further assess the relationship between EC metrics and behavioral symptoms, we also adopted a data-driven dimensionality reduction approach. Specifically, we performed a principal component analysis (PCA) using three principal components to predict SRS scores separately in ASD and TD groups (Supplementary Figure 3), comparing models based on normative EC metrics and standard EC metrics. The number of principal components was chosen as the minimum number capable of explaining 60% of the observed variance in all SRS subscales.

Prediction performance was higher when using normative EC metrics in both groups. In ASD, normative metrics explained a larger proportion of variance in SRS Total T-scores compared with standard EC metrics ($R^2$=0.49 vs $R^2$=0.33), and a similar pattern was observed in TD ($R^2$=0.44 vs $R^2$=0.24).To test the robustness of this effect, the analysis was repeated for all SRS subscales, comparing the resulting $R^2$ values obtained with normative versus standard EC metrics. In both ASD and TD groups, normative EC metrics consistently provided significantly better predictions (paired t-test: t=-4.63, p=0.002, d=1.54 for ASD; t=-4.32, p=0.003, d=1.44 for TD). A further analysis on Symptom-specific association of Normative EC metrics can be found in Supplementary Material.

##### Symptom-specific association of Normative EC metrics

We investigated the association between EC metrics and the different SRS subscales. For each subscale, we retained only correlations exceeding a predefined threshold ($R^2$>0.42, the threshold value was chosen to allow for a statistical comparison between different EC metrics, see Methods) and characterized the resulting set of connections in terms of EC metric families and topographical properties of the channels involved.

For Social Communication and Interaction (SCI, Supplementary Figure 4a), 15 EC metrics exceeded the threshold. The most represented family was dDTF (11 metrics), and the distribution across EC metric families significantly deviated from chance ($\chi^2$ test, p=0.00002). Topographically, connections showed a significant predominance of frontal sources (p=0.0003) and right-hemisphere sources (p=0.007). A significant bias was also observed for frontal targets (p=0.028) and right-hemisphere targets (p=0.0001).

For Social Awareness (AWR, Supplementary Figure 4b), 19 EC metrics exceeded the threshold. The most represented family was PDC (12 metrics), with a significant difference across EC metric families ($\chi^2$ test, p=0.008). All connections originated from frontal channels and terminated in frontal channels (p=0.007), while no significant lateralization effects were observed for either sources (p=0.17) or targets (p=0.17).

For Social Cognition (COG, Supplementary Figure 4c), 7 EC metrics exceeded the threshold. The most represented family was dDTF (3 metrics), but the distribution across EC metric families did not significantly differ from chance ($\chi^2$ test, p=0.27). No significant topographical biases were observed for connection sources or targets (all p=0.23).

For Restricted Interests and Repetitive Behaviours (RRB, Supplementary Figure 4d), 5 EC metrics exceeded the threshold. The most represented family was dDTF (3 metrics), and the distribution across EC metric families was not significantly different from chance ($\chi^2$ test, p=0.092). A modest predominance of frontal sources was observed (p=0.045), while no other topographical effects were significant (all p≥0.18).

For Social Communication (COM, Supplementary Figure 4e), 11 EC metrics exceeded the threshold. The most represented family was dDTF (8 metrics), and the distribution across EC metric families significantly differed from chance ($\chi^2$ test, p=0.0004). While no significant bias was observed for connection sources (frontal p=0.050, right p=0.23) or frontal targets (p=0.11), a significant predominance of right-hemisphere targets was found (p=0.0001).

For Social Motivation (MOT, Supplementary Figure 4f), 11 EC metrics exceeded the threshold. The most represented family was DTF (6 metrics), and the distribution across EC metric families significantly differed from chance ($\chi^2$ test, p=0.024). Connections preferentially originated from frontal channels (p<0.00001) and terminated in frontal channels (p=0.020), while no significant lateralization effects were detected (sources p=0.23, targets p=0.23).

For completeness, we also examined the SRS total score (TOT, Supplementary Figure 5), for which 11 EC metrics exceeded the threshold. The most represented family was dDTF (5 metrics), with a marginally non-significant difference across EC metric families ($\chi^2$ test, p=0.051). No significant bias was observed for connection sources (frontal p=0.050, right p=0.99), whereas targets showed a significant predominance of frontal (p=0.020) and right-hemisphere channels (p=0.012).

**Theta and Alpha Effective Connectivity**

Barnes et al. (26) reported results consistent to our findings using wavelet transfer function EEG analysis. Although their study focused only on ASD–TD group differences and did not investigate clinical severity, they reported significant alterations in frontal theta and alpha connectivity, which were also confirmed in our cohort. In our analysis, similarly, the strongest ASD–TD difference was observed in ffDTF of theta band from Fp1 to Fp2 (ffDTF $^{q}{}_{Fp1\text{->}Fp2}$) using Mann–Whitney testing after FDR-BH correction (q = 0.002, Cohen's d = 0.78), and was further supported by logistic regression controlling for age and sex (p = 0.0002), while age and sex themselves were not significant predictors ($p_{AGE}$ = 0.64; $p_{SEX}$ = 0.31).

More in general, we can summarize our findings stating that dysconnectivity in lower bands (delta, theta) of the relevant frontal electrodes show a group difference from ASD subjects respect to TD subjects which confirms previous findings (26) and leads to the classification previously discussed (see results); given this, clinical severity increases with an increasing weighted normative deviation in the tempo-parietal and frontal electrodes (see Figure 4b right panel).

**Robustness Analysis of EC metrics**

We further assessed robustness through an additional validation analysis performed on 10 matched pairs of subjects ASD–TD (20 subjects total), using the raw EC values prior to normative modeling. Two complementary sources of methodological variability were evaluated: (i) reduction of the MVAR model order and (ii) intra-subject variability across different non-overlapping 10-s resting-state segments. Robustness was quantified using Pearson correlation between the reference EC matrices (baseline configuration: model order = 20, representative segment) and the EC matrices obtained under each alternative condition. Group-level robustness was quantified using the mean Pearson correlation coefficient across matched subjects, with higher correlation values indicating greater stability of the EC structure across methodological variations.

For model order sensitivity, the baseline configuration (order = 20) was compared with progressively lower orders (15, 10, and 6). Pearson correlation remained high across all comparisons ($r = 0.90$ for order 15, $r = 0.82$ for order 10, and $r = 0.79$ for order 6), indicating that the EC structure was substantially preserved even under reduced autoregressive complexity. Intra-subject robustness was then assessed by comparing EC estimates obtained from different resting-state segments of the same subject. Pearson correlation remained high ($r \approx 0.80$), indicating good stability of subject-specific EC profiles across segments (see Supplementary Figure 8). These results support the reliability of the selected EC metric and justify the use of both the representative segment and the fixed model order adopted in the main analyses.

#### Supplementary Tables

##### Table S1

| | Number of participants | |
|---|---|---|
| | TD | ASD |
| Center 1 | 66 (48) | 35 (30) |
| Center 2 | 12 (11) | 21 (18) |
| Center 3 | 0 | 15 (10) |

**Table S1. Participants numbers in different clinics:** Participant number are reported for different groups, male numbers are reported in brackets.

**Table S2.**

| SRS subscale | Mean TD | Std TD | Mean ASD | Std ASD | stat | p-val | p-site | p-interaction |
|---|---|---|---|---|---|---|---|---|
| Age | 10.0 | 3.4 | 10.2 | 3.5 | 2664.0 | 0.69 | 0.21 | 0.61 |
| SRS - SCI | 50.8 | 9.3 | 72.0 | 13.0 | 525.5 | <0.00001 | 0.65 | 0.37 |
| SRS - AWR | 52.2 | 8.3 | 68.9 | 11.7 | 656.5 | <0.00001 | 0.93 | 0.072 |
| SRS - COG | 49.8 | 9.7 | 70.9 | 13.4 | 590.0 | <0.00001 | 0.97 | 0.29 |
| SRS - RRB | 49.1 | 8.4 | 73.7 | 12.4 | 279.0 | <0.00001 | 0.43 | 0.50 |
| SRS - COM | 50.5 | 9.0 | 71.8 | 13.2 | 538.0 | <0.00001 | 0.57 | 0.65 |
| SRS - MOT | 50.4 | 9.8 | 65.4 | 13.6 | 1004.0 | <0.00001 | 0.41 | 0.058 |
| SRS – TOT | 50.0 | 9.2 | 72.9 | 13.0 | 434.0 | <0.00001 | 0.69 | 0.43 |

**Table S2: Age values and SRS subscale values across groups.** P-value for differences across sites, and for the interaction between sites and diagnostic groups are also reported.

**Table S3.**

| SRS subscale | 10-fold $R^2$ | p-value | $R^2$ (EC \| SRS) |
|---|---|---|---|
| SCI | 0.24 | 0.0004 | 0.04 |
| AWR | 0.09 | 0.004 | 0.02 |
| COG | 0.09 | 0.038 | 0.01 |
| RRB | 0.004 | 0.003 | 0.004 |
| COM | 0.27 | 0.0004 | 0.03 |
| MOT | 0.12 | 0.006 | 0.002 |
| Total | 0.23 | 0.0005 | 0.03 |

**Table S3: Regression coefficients and p-values for SRS subscales.** Results are reported after 10-fold cross-validation. Partial $R^2$ values after correction for SRS subscales intercorrelations are reported in the rightmost columns, p-values (all $p>0.05$) are not reported.

**Table S4.**

| EC metrics | Band | Input | Target | SRS explained variance |
|---|---|---|---|---|
| dDTF08 | Beta | TP8 | Fp2 | 61.9 % |
| dDTF08 | Gamma | TP8 | Fp2 | 14.6 |
| dDTF08 | Beta | TP8 | F4 | 10.9 |
| dDTF08 | Gamma | TP8 | F4 | 9.9 |
| dDTF08 | Beta | Fp1 | Fp2 | 1.9 |
| dDTF08 | Gamma | Fp1 | Fp2 | 0.4 |
| dDTF08 | Beta | Fp2 | Fp1 | 0.3 |
| dDTF08 | Beta | Fp2 | F3 | 0.1 |

**Table S4: Selected normative EC metrics for SRS prediction.** All metrics reported refer to their normative value. It can be appreciated that they are all Right Tempo-Parietal to Right Frontal electrodes in beta & gamma, as well as Frontal electrodes in beta & gamma. Explained variance for SRS Total is also reported in the rightmost column.

**Table S5.**

| EC | Regression Coefficient ASD | Regression Coefficient TD |
|---|---|---|
| Beta $dDTF08_{TP8 \rightarrow Fp2}$ | 9.585 | 3.331 |
| Gamma $dDTF08_{TP8 \rightarrow Fp2}$ | 0.774 | 3.809 |
| Beta $dDTF08_{TP8 \rightarrow F4}$ | -2.976 | -1.757 |
| Gamma $dDTF08_{TP8 \rightarrow F4}$ | 1.396 | -3.989 |
| Beta $dDTF08_{Fp1 \rightarrow Fp2}$ | 0.215 | 1.707 |
| Gamma $dDTF08_{Fp1 \rightarrow Fp2}$ | -3.023 | -1.610 |
| Beta $dDTF08_{Fp2 \rightarrow Fp1}$ | -2.323 | 1.127 |
| Beta $dDTF08_{Fp2 \rightarrow F3}$ | -0.431 | -2.782 |
| Intercept | 74.364 | 49.904 |

**Table S5: Regression coefficients for SRS Total scale.**

**Table S6.**

| Age Stratum | 5 – 7 | 7 - 10 | 10 - 13 | 13 - 20 | Total |
|---|---|---|---|---|---|
| $N_{ASD}$, $N_{TD}$ | 10, 10 | 18, 19 | 14, 15 | 15, 13 | 57, 57 |
| $ASD_{Age}$ (mean ± SD) | 5.712 ± 0.406 | 8.096 ± 0.823 | 11.310 ± 0.737 | 14.743 ± 1.675 | 10.216 ± 3.432 |
| $TD_{Age}$ (mean ± SD) | 5.646 ± 0.486 | 8.012 ± 0.709 | 11.408 ± 0.683 | 14.452 ± 0.910 | 9.959 ± 3.194 |
| %Female (ASD, TD) | 30.0, 30.0 | 0.0, 5.3 | 14.3, 20.0 | 26.7, 38.5 | 15.8, 21.1 |
| SMD Age | 0.147 | 0.109 | -0.138 | 0.216 | 0.078 |
| SMD Sex | 0.000 | -0.329 | -0.152 | -0.252 | -0.136 |

**Table S6: Demographic characteristics of the final analytical subset stratified by age.** Demographic characteristics of the final analytical subset (57 ASD and 57 TD participants) stratified by age. At the whole-group level, no significant between-group differences were observed for age (Welch's t-test, $p = 0.68$) or sex distribution ($\chi^2$ test, $p = 0.47$). Standardized mean differences (SMD) are reported for age and sex within each predefined age stratum.

### Supplementary Figures

#### Figure S1.

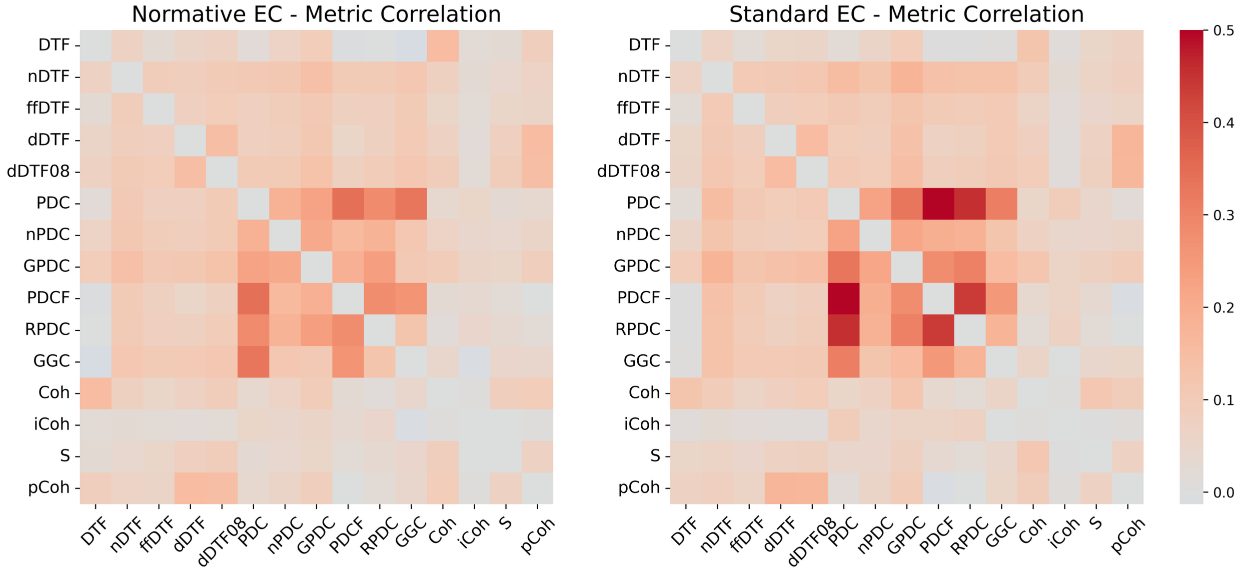


**Figure S1: Correlation matrices:** Correlation matrices between normative EC metrics (left) and standard EC metrics (right).

#### Figure S2.

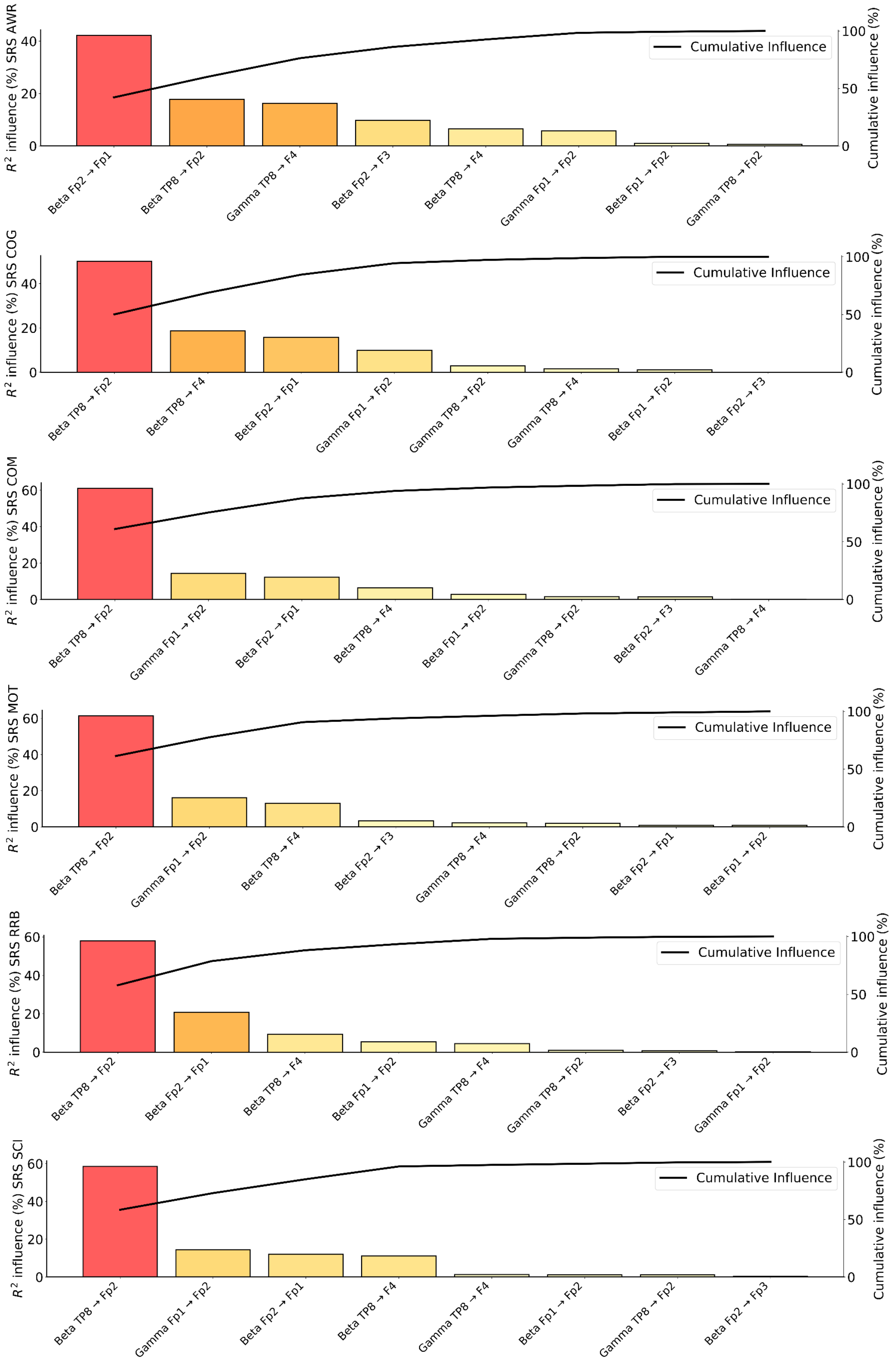


**Figure S2: Pareto chart showing the influence of each EC quantity on the different SRS subscales (a):** AWR subscale. **(b):** COG subscale. **(c):** COM subscale. **(d):** MOT subscale. **(e):** RRB subscale. **(f):** SCI subscale.

#### Figure S3.

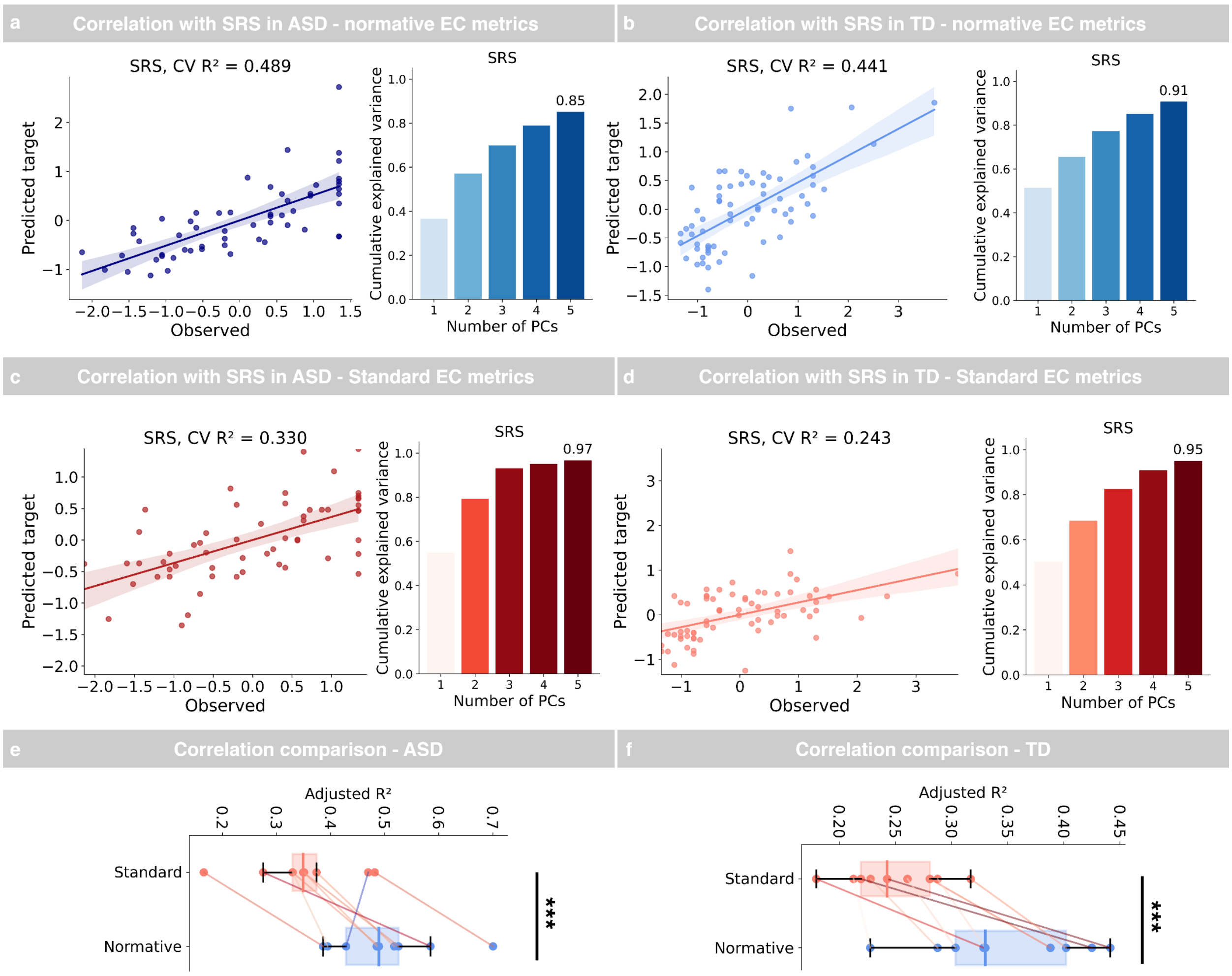


**Figure S3: PCA prediction of SRS Total based on normative EC metrics. (a):** Linear regression between ASD SRS Total T values and top 3 PCAs from normative EC metrics (left), with variance explained by top 5 PCAs (right). **(b):** Linear regression between TD SRS Total T values and top 3 PCAs from normative EC metrics (left), with variance explained by top 5 PCAs (right). **(c):** Linear regression between ASD SRS Total T values and top 3 PCAs from standard EC metrics (left), with variance explained by top 5 PCAs (right). **(d):** Linear regression between TD SRS Total T values and top 3 PCAs from standard EC metrics (left), with variance explained by top 5 PCAs (right). **(e):** Boxplot comparing $R^2$ values of the linear regressions between SRS subscales and standard EC (red) and normative EC (blue) metrics for the ASD group. **(f):** Boxplot comparing $R^2$ values of the linear regressions between SRS subscales and standard EC (red) or normative EC (blue) metrics for the TD group. Significance notation: *** stands for p<0.0005.

#### Figure S4

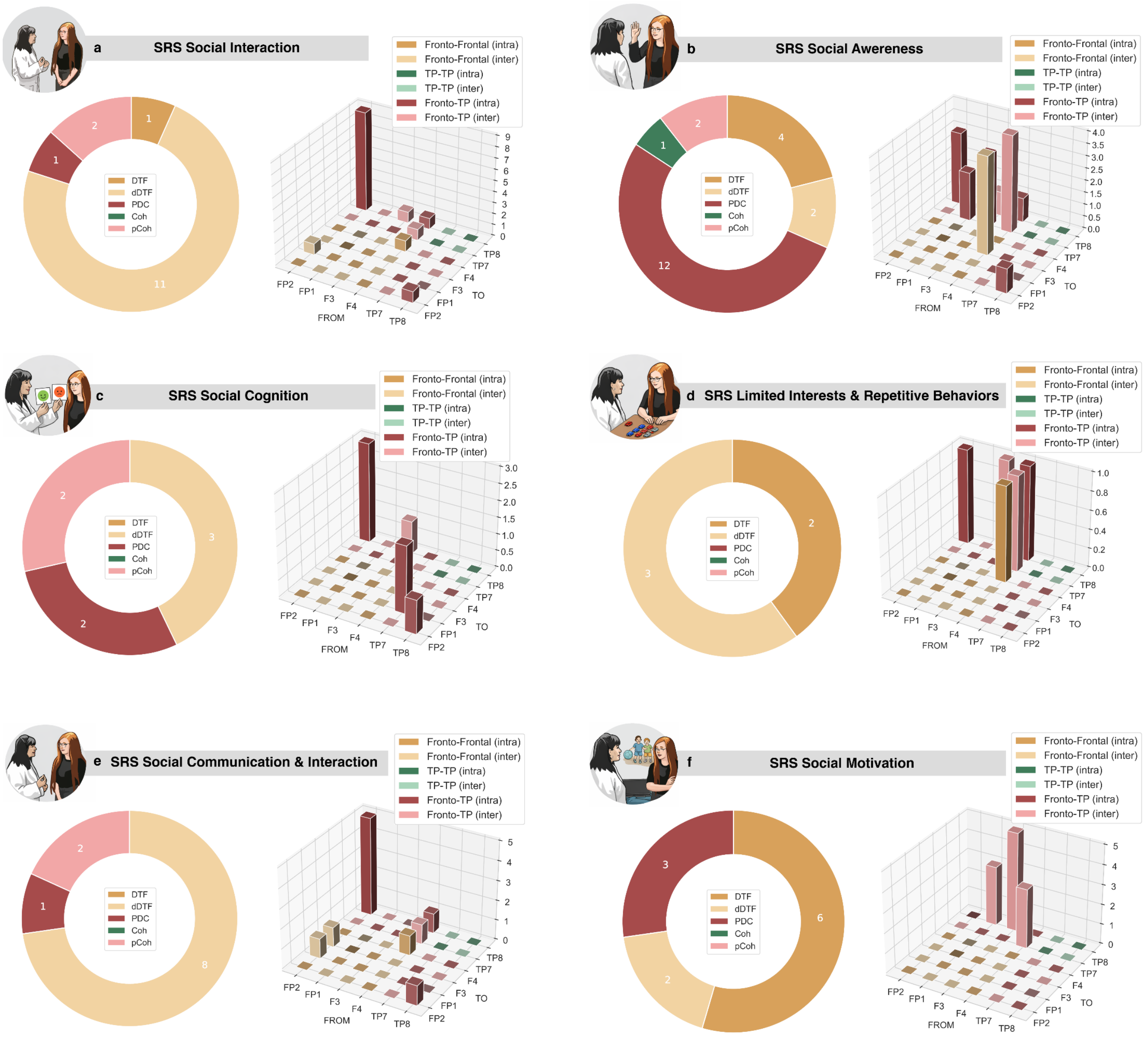


**Figure S4: Symptom-specific association of normative EC metrics. (a):** Pie-chart reporting the family of normative EC metrics significatively associated with SRS - Social Interaction subscale (left). Bar-plot reporting the topography of normative EC metrics significatively associated with SRS - Social Interaction subscale (right). **(b):** Family pie-chart and topography bar-plot for SRS - Social Awareness subscale. **(c):** Family pie-chart and topography bar-plot for SRS - Social Cognitive subscale. **(d):** Family pie-chart and topography bar-plot for SRS - Repetitive Behaviors. **(e):** Family pie-chart and topography bar-plot for SRS - Social Communication subscale. **(f):** Family pie-chart and topography bar-plot for SRS - Social Motivation subscale.

**Figure S5.**

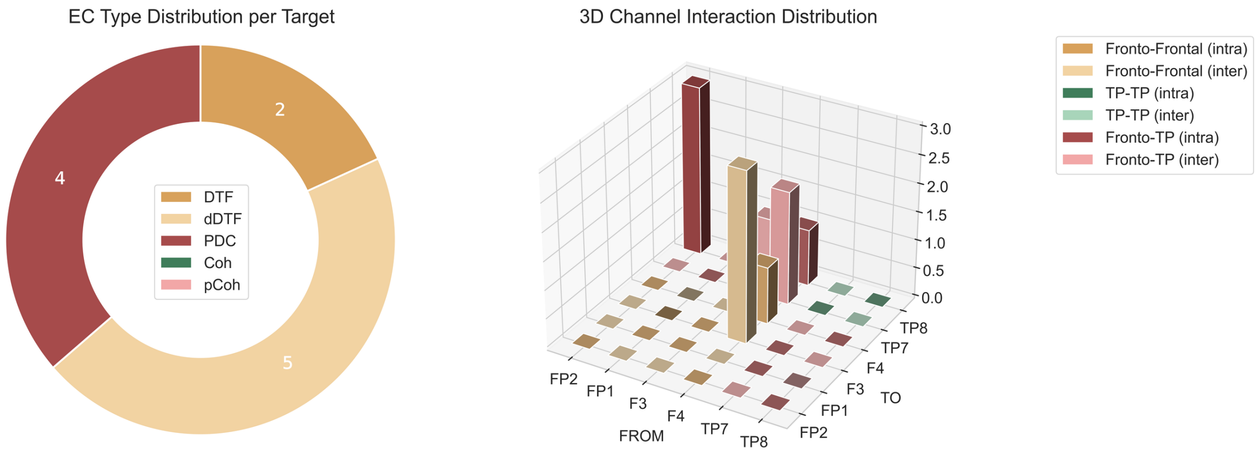


**Figure S5: Association of normative EC metrics with SRS Total:** Pie-chart reporting the family of normative EC metrics significatively associated with SRS - Total (left). Bar-plot reporting the topography of normative EC metrics significatively associated with SRS - Total (right).

**Figure S6**

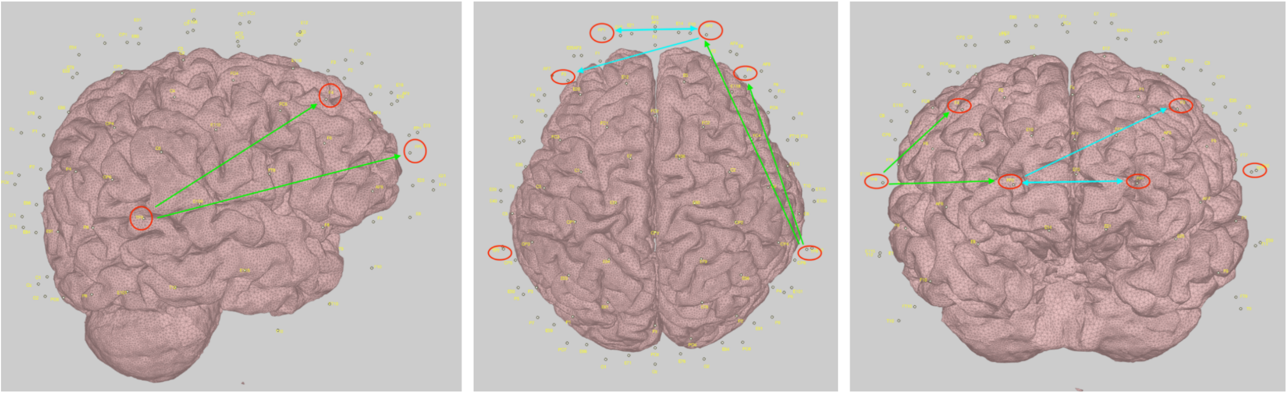

**Figure S6: Position of electrodes considered for regression in ASD symptom severity**

### Figure S7.

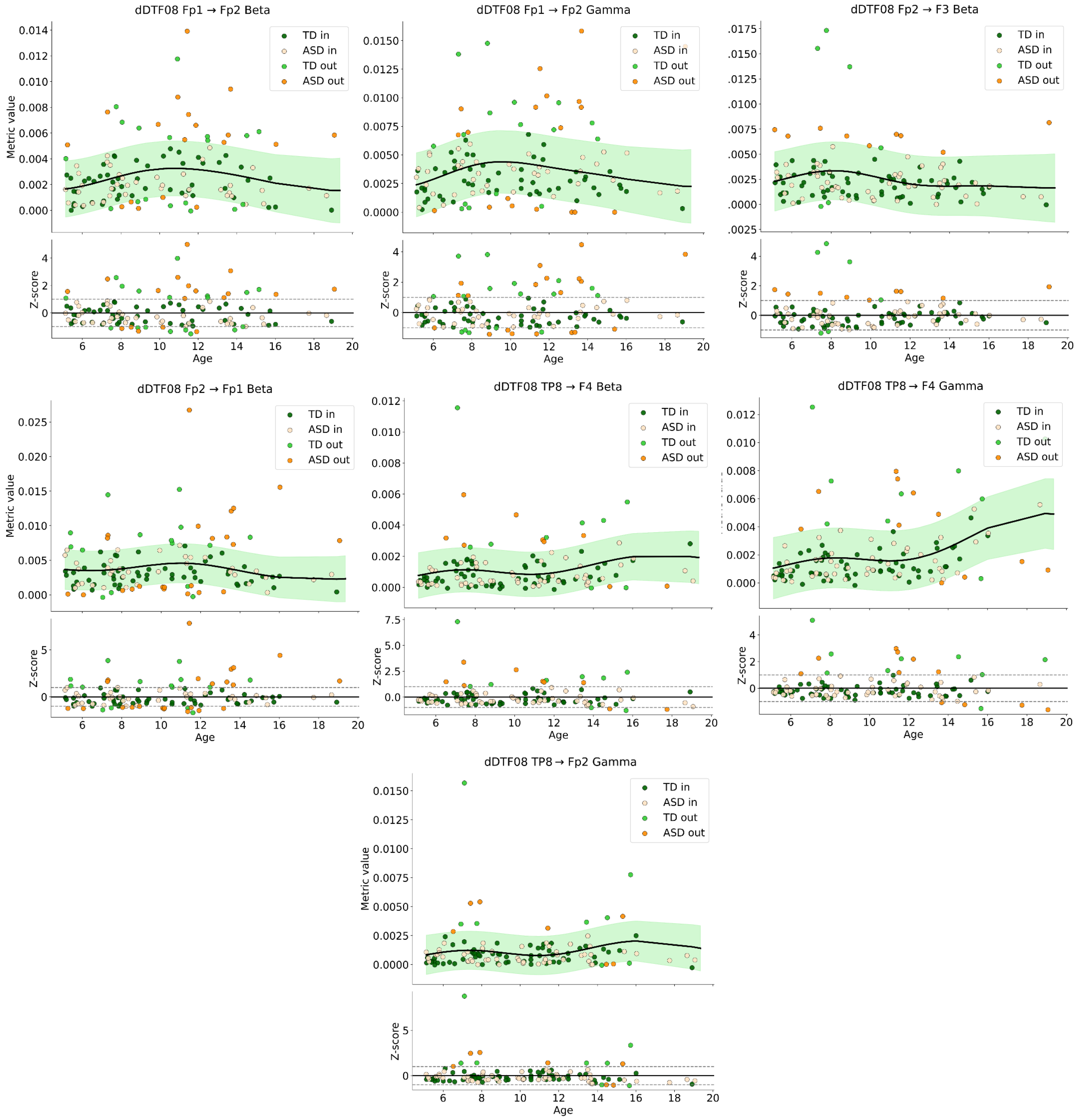


**Figure S7: Normative curves for the selected EC measures.** Each panel represents the normative trajectory of each of the selected EC feature estimated using Gaussian Process Regression (GPR) on the typically developing (TD) group. The black line represents the age-dependent normative mean, the shaded area indicates the normative range (±2 standard deviations), and dots represent observed EC values. Normative residuals are also reported for each metric. Dashed horizontal lines indicate the ±2 z-scores. Positive and negative values indicate hyperconnectivity and hypoconnectivity relative to the normative developmental trajectory, respectively.

**Figure S8.**

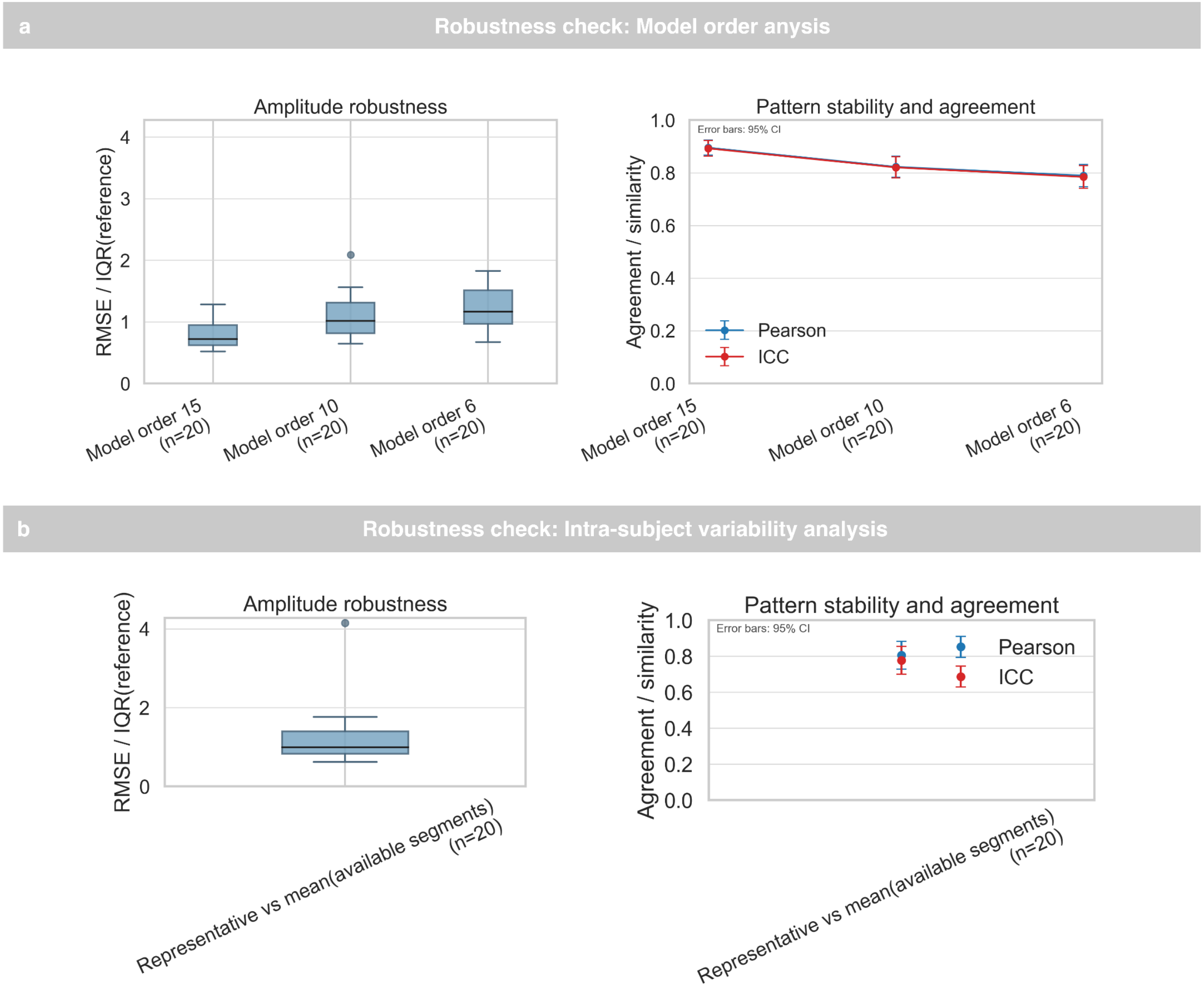


**Figure S8: Robustness analysis of Effective Connectivity. (a):** Stability of EC estimates under changes in MVAR model order. Pearson correlation was computed between the baseline configuration (model order = 20) and progressively lower model orders (15, 10, and 6), showing high preservation of the connectivity structure across configurations (r = 0.90, 0.82, and 0.79, respectively). **(b)** Intra-subject variability analysis. Pearson correlation between EC matrices derived from different non-overlapping 10-s resting-state segments of the same subject remained high (r ≈ 0.80), indicating good stability of subject-specific EC profiles across segments. These results support the robustness of the selected EC metric and justify the use of a fixed model order and a representative segment in the main analyses.
